# Long-term intrinsic cycling in human life-course antibody responses to influenza A(H3N2)

**DOI:** 10.1101/2022.06.27.22276898

**Authors:** Bingyi Yang, Bernardo Garcia-Carreras, Justin Lessler, Jonathan M. Read, Huachen Zhu, C. Jessica E. Metcalf, James A. Hay, Kin On Kwok, Ruiyin Shen, Chao Qiang Jiang, Yi Guan, Steven Riley, Derek A.T. Cummings

## Abstract

Over a life-course, human adaptive immunity to antigenically mutable pathogens exhibits competitive and facilitative interactions. We hypothesize that such interactions may lead to cyclic dynamics in immune responses over a lifetime. Here, we demonstrated a long-term periodicity (about 24 years) in individual antibody responses, by analyzing hemagglutination inhibition titers against 21 historical influenza A(H3N2) strains spanning 47 years from a cohort in Guangzhou, China. The reported cycles were robust to analytic and sampling approaches. Simulations suggested that individual-level cross-reaction between antigenically similar strains likely explain the reported cycle. We showed that the reported cycles are predictable at both individual and birth-cohort level and that cohorts show a diversity of phases of these cycles. Phase of cycle was associated with the risk of response to circulating strains, after accounting for age and pre-existing titers of the circulating strains. Our findings reveal the existence of long-term periodicities in individual antibody responses to A(H3N2). We hypothesize that these cycles are driven by pre-existing antibody responses blunting responses to antigenically similar pathogens (by preventing infection and/or robust antibody responses upon infection), leading to reductions in antigen specific responses over time until individual’s increasing risk leads to an infection with an antigenically distant enough virus to generate a robust immune response. These findings could help disentangle cohort-effects from individual-level exposure histories, improve our understanding of observed heterogeneous antibody responses to immunizations, and inform targeted vaccine strategy.

## MAIN

Over a life-course, a key feature of human adaptive immune responses is the ability to continually update and refine responses to new antigens. A key example is immune responses to influenza, a pathogen that is constantly experiencing genetic and antigenic change. Antibodies mounted against a specific influenza virus decay after exposure until re-exposure or infection to an antigenically similar virus occurs, whereupon back-boosting of antibodies acquired from previous infections can occur, as well as updating antigen specific antibodies to the newly encountered infection (Amanna et al., 2007; Edridge et al., 2020; Fonville et al., 2014; Kucharski et al., 2018). As antibodies are considered a correlate of protection from infection (Cowling et al., 2019; Dunning, 2006; Krammer, 2019; Truelove et al., 2016), studies often measure antibodies against the circulating strain to estimate risk of infection. However, interactions between antibodies that were acquired from recent and long-ago infections can mean that characterization of antibodies to only currently circulating strains of pathogens may only partially capture antibody protection and risk of infection (Cowling et al., 2019; Ng et al., 2019; Yang et al., 2020).

Antibody mediated immune response to multiple infections generated through repeated exposures to antigenically variable pathogens result in not only the facilitative interactions (e.g., back-boosting and cross-protection (Krammer, 2019)), but also competitive interactions (e.g., immune imprinting (Gostic et al., 2016; Reynolds et al., 2022; Vieira et al., 2021), antigenic seniority (Lessler et al., 2012)). Non-neutralizing antibodies acquired from previous exposures can be boosted (Gouma et al., 2020) and may blunt the immune responses to new infections. Immune functions targeting antigenically specific pathogens may rise or lower in prevalence over a person’s lifetime, in response to both a new infection and these competitive and facilitative interactions. Such interactions provide positive and negative feedbacks that have routinely been found to drive cycles in other systems (e.g., predator-prey, host-parasite) (Post and Palkovacs, 2009; Yoshida et al., 2003). Therefore, we might expect feedback mechanisms to introduce intrinsic temporal cycles in an individual’s immune responses to antigenically variable pathogens over a lifetime, yet these cycles have not often been investigated.

Here, we examine seasonal influenza as a case study. Three subtypes of influenza (A(H3N2), A(H1N1) and B) cause an estimated 291,000-645,000 deaths globally every year (Iuliano et al., 2018). Although viruses of the same subtype share similar surface proteins, continuous genetic mutation leads to antigenic variation, resulting in escape from immune recognition by antibodies generated by previous infections. However, escape is not complete. Cross-reactive immunity across strains exists for viruses isolated at different times(Bedford et al., 2014; Fonville et al., 2014; Krammer, 2019; Smith et al., 2004). While high levels of antibody have been found to be protective from infection, they have also been found to be associated with reduced antibody responses to new infections and influenza vaccination (Auladell et al., 2022). New infections were found to boost antibodies against previously encountered viruses as much if not more than that of the infecting virus(Auladell et al., 2022). Therefore, we hypothesized that the combination of antigen specific and non-specific responses may give rise to cycles in antibody responses over an individual’s life span. We tested the hypothesis that human adaptive immune responses exhibit nonlinear interactions with evolving viruses, creating intrinsic cycles in antibody responses.

To test the hypothesis, we characterized the periodic behavior of 777 paired antibody profiles, measured in 2010 (baseline) and 2014 (follow-up), measuring antibody responses (i.e., hemagglutination inhibition (HI) titers) to 21 A(H3N2) strains circulating over 47 years (Fig. 1A and Fig. S1A and S2) (Jiang et al., 2017; Yang et al., 2020). We used Fourier analysis to examine the periodicity of individual antibody responses, after accounting for shared variations arising from virus-specific population-level circulation and/or laboratory measurement. We assessed the robustness of the observed cycles to multiple analytic and sampling methods. We then used a previously published mechanistic model that characterizes individual antibody responses to a set of antigenically similar strains to test the sensitivity of these cycles to multiple generally recognized biological mechanisms (Kucharski et al., 2018). Finally, we determined whether the cyclic pattern in individual antibody responses is predictable and whether it could improve the prediction of the risk of seroconversion to circulating strains of influenza A(H3N2) over existing models.

**Figure 1.**
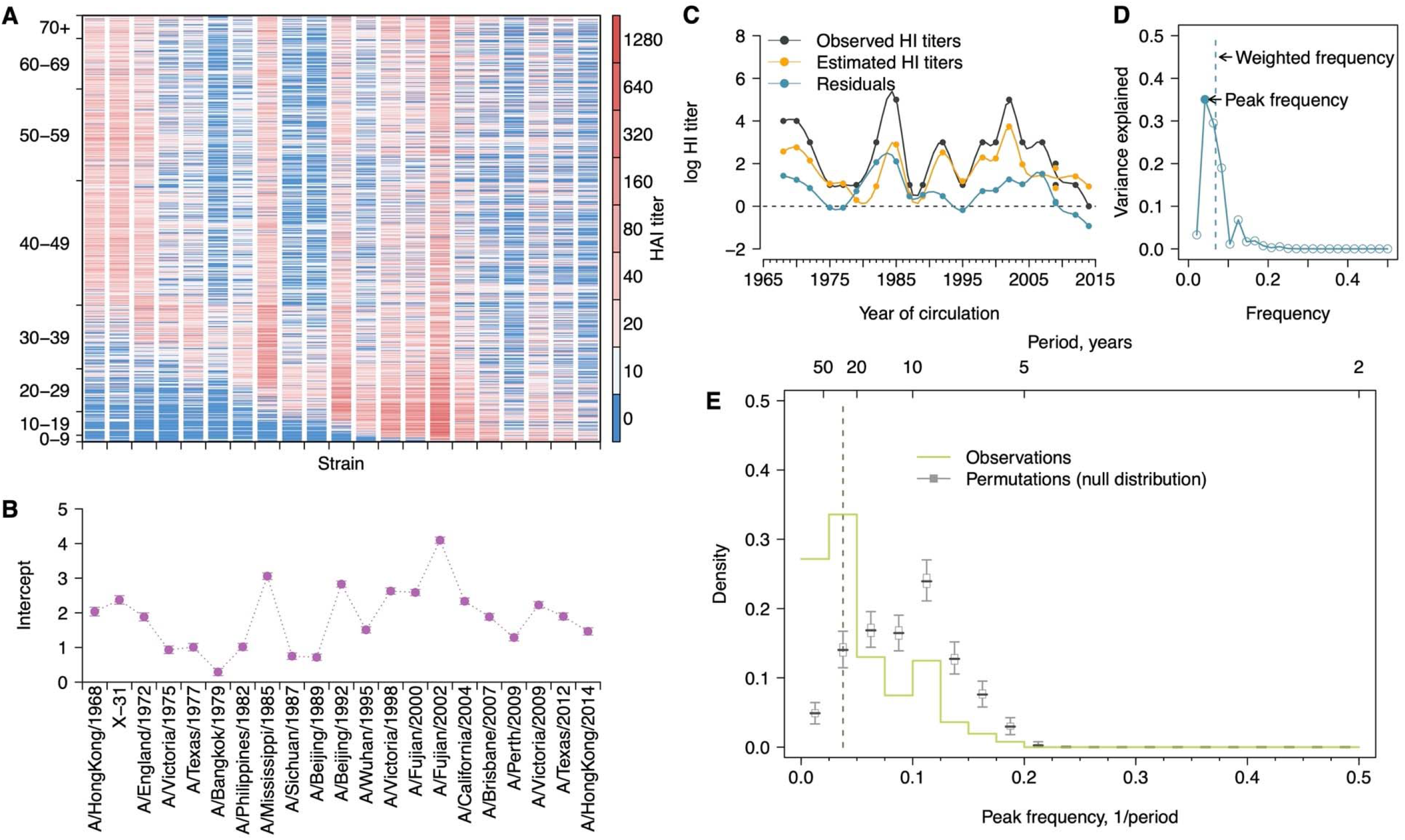
Long-term cycles in individual antibody responses to influenza A(H3N2) at baseline. **(A)** HI titers against A(H3N2) strains at baseline. Each row shows an antibody profile for a participant. Participants are sorted by age (y-axis). Strains (x-axis) are sorted by the year of isolation, which are listed in the x-axis of panel B. **(B)** Strain-specific intercepts. A generalized additive model (GAM) was fitted to log HI titers (shown in A) on age at sampling (spline), age at isolation (spline) and strains (categorical) (also used for C). With the model, we extracted strain-specific intercepts (representing population level activity; shown in B) and calculated the residuals between predicted and observed log HI titers for each individual (individual level antibody responses; shown in C and used for D-E. Details in Fig. S4A). (C) Illustration of estimating individual time series of residuals. Estimates were derived from the GAM model in panel B. Residuals were calculated as the difference between observed and estimated HI titers (i.e., black minus orange; shown as the blue line). **(D)** Illustration of a Fourier spectrum. Peak (i.e., the frequency explaining the largest variance) and weighted frequency of a Fourier spectrum of the interpolated time series of residuals shown in C. **(E)** Distribution of peak frequencies of individual residuals. We performed Fourier spectral analysis (shown in D) on the time series of residuals of each person and extracted the peak frequency. The light green shows the distribution of peak frequencies across participants, with the dashed vertical line indicate the peak frequencies that had the highest proportions among individuals. Median (thick grey ticks), interquartile (grey boxes) and 95% intervals (thin grey ticks) of distributions from 1,000 permutations.

## Results

### Identifying long-term cycles in individual antibody responses to influenza A(H3N2)

Antibody titers against a set of strains isolated over 47 years, when ordered by the time of isolation of the tested strains, form a time series that describes the immune history of an individual (Fig. S3) (Yang et al., 2020). To describe variations in these time series attributable to virus specific and/or individual-level host characteristics, we fitted a generalized additive model (GAM) of log-titers with strain-specific intercepts and non-linear effects of age at serum collection (i.e., biological age) and age at the year when strains were isolated (i.e., birth-cohort effect) (Kucharski et al., 2018). Strain-specific intercepts (Fig. 1B and Fig. S1B) were estimated to adjust for the average population antibody responses due to A(H3N2) circulation and/or virus specific differences in laboratory assay measurements. Residuals were then estimated to represent individual-level departures from population averages (Fig. 1C and S4) and were interpolated to annual resolution with spline function (see details in Methods).

We investigated whether cyclic behavior was present in antibody responses by performing Fourier analysis on each individual’s time series of residuals (Fig. S4). The periodicity for each participant was determined by the frequency (“peak frequency” hereafter) that explained the most variance in the Fourier spectrum (Fig. 1D). To test the significance of these peak frequencies, we compared the distribution of peak frequency across participants with those distributions (i.e., null expectation) from 1,000 permutations, in which observations for each time series were shuffled (Fig. S5C-D). This null expectation represents the peak frequency distribution of random non-periodic time series and reflects the underlying structure that is introduced by our sampling and interpolation approaches (Fig. S6-7).

We found 33.6% (95% CI, 30.3% to 37.0%) of participants had a peak frequency corresponding to a long-term periodicity (i.e., 20 to 40 years, translated from frequencies of 0.025 to 0.050; see Methods) at baseline, which was significantly higher than null expectation (Fig. 1E and Fig. S1C), suggesting that the observed cyclic patterns were not purely due to chance. This peak frequency range (corresponded to a ∼24-year periodicity) accounted for a median 22.1% of the variance [interquartile range (IQR), 11.1% to 35.4%] of individual-level residuals (Table S1).

We conducted multiple sensitivity analyses and validations to test the robustness of the observed cycles in individual-level antibody responses to analytic methods and our sampling methods. Across these analyses, including methods that accounted for variation in each individual’s spectra (Fig. 2A), irregularity in isolation intervals of tested strains (Fig. 2B) and secular trends in our time series (Fig. 2C and Fig. S8), we found consistent evidence for long-term periodicity in antibody responses. Results were robust to leaving specific strains out of the analysis (Fig. S9-10) and method of interpolation (Fig. S6-7). A full description of sensitivity analyses including validation in subsets of our data is provided in Methods (Fig. S6-11).

**Figure 2.**
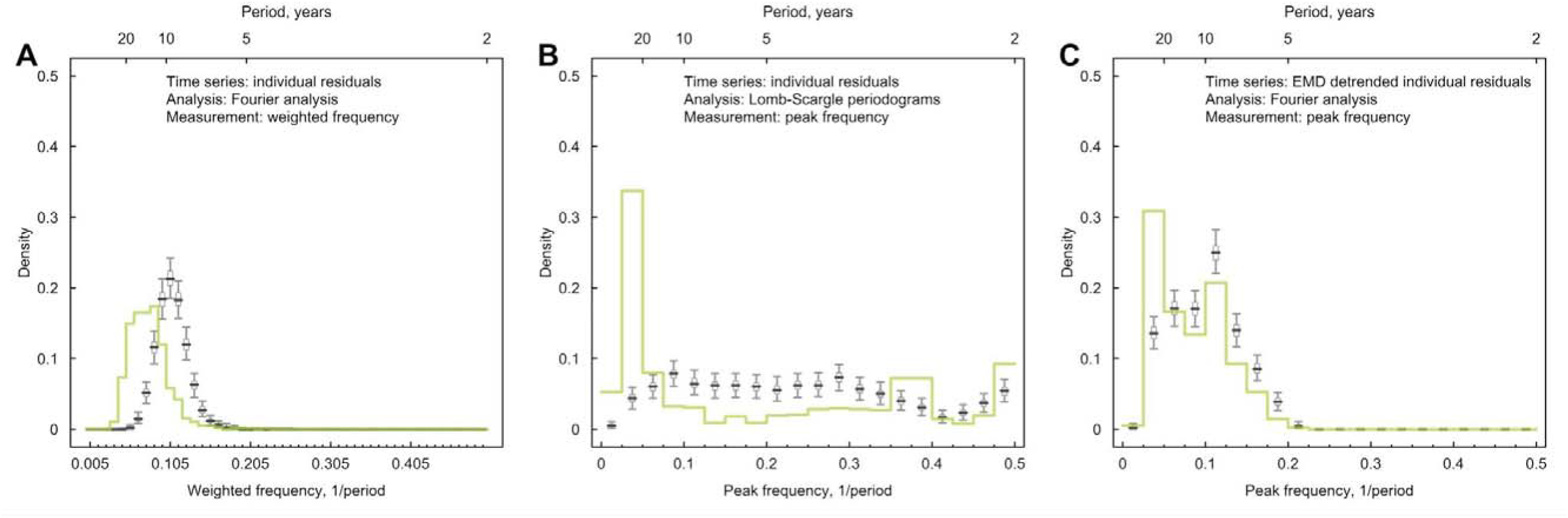
Impacts of irregularly sampled data, interpolations and long-term trends on cycles identified in individual antibody responses at baseline. **(A)** Distribution of weighted frequencies of individual Fourier spectra at baseline. We performed Fourier spectral analysis on the interpolated time series of residuals for each person and calculated the average frequency weighted by the variance explained (‘weighted frequency’; see Fig. 1D). **(B)** Distribution of peak frequencies of individual Lomb-Scargle periodograms. We performed Lomb-Scargle periodograms on the time series of residuals for each person and extracted the frequency that explained the most variance (‘peak frequency’). **(C)** Distribution of peak frequencies of individual Fourier spectra of detrended residuals at baseline. We performed Fourier analysis on time series that removed the non-linear trend identified using empirical mode decomposition (EMD) analysis.

Additionally, we analyzed an independent out-of-sample data set from Vietnam (HI titers of 57 strains for 69 participants measured annually, 2007-2012 (Horby et al., 2012; Kucharski et al., 2018)). Due to the lack of data on age, we compared long-term periodicity in HI titers and found a similar long-term periodicity in both studies (Fig. S12), suggesting that similar cycling is likely present in other settings, even with population-level variations.

### Cycles in individual antibody responses likely associated with homotypic cross-immunity

To investigate possible biological mechanisms, we simulated individual antibody profiles encompassing known feedbacks and interactions due to generally recognized immunological mechanisms (Table S2-3). We primarily applied a model by Kucharski *et al*. ((Kucharski et al., 2015)) that describes the snapshot of individual antibody dynamics, resulting from varied individual infection histories, narrow (i.e., against recent strains) and broad (i.e., against distant strains) range of cross-reactions of antigenically similar strains and antibody waning (Figure S14; *Equation 9*). We extended the model to allow for the influence of individual-level pre-existing antibodies and population-level viral activity on individual infection hazard (Fig. 3I and S13; *Equations 10-11*). Infection events are simulated annually and individually according to individual infection hazard, which is then used to inform the updated antibody profiles using Kucharski’s model (Fig. S13). As viral circulating pattern at population-level is not the focus of this study and its potential drivers (e.g., arising from homotypic and/or heterotypic cross-immunity) are inconclusive, we therefore assumed two scenarios (Fig. 3I) to examine the impact of predictable (i.e., cyclic) or non-predictable (i.e., random) annual attack rates on the observed individual antibody responses.

**Figure. 3.**
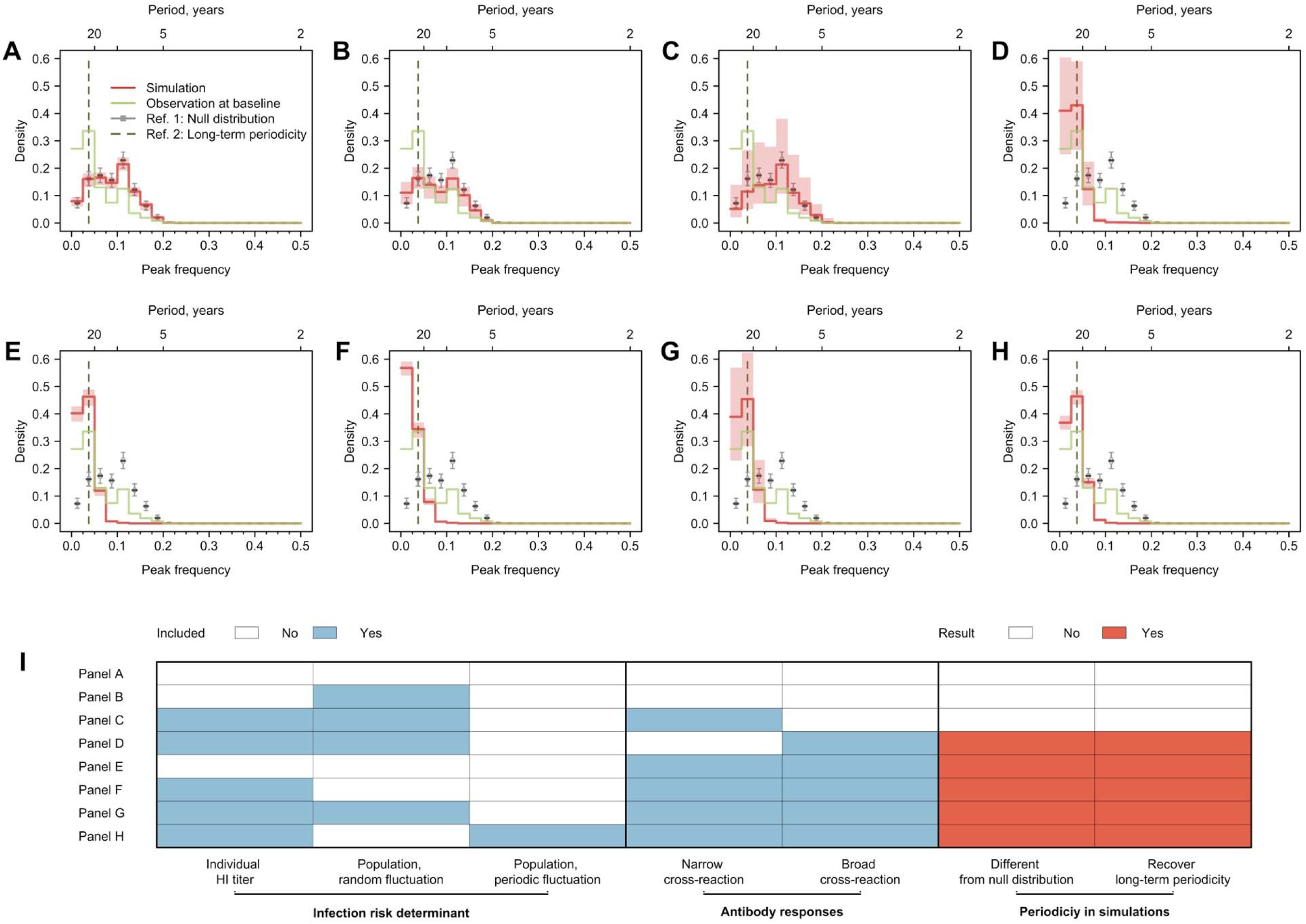
Cycles in simulated antibody responses from the model accounted for different mechanisms. Colored lines are the distribution of peak frequencies detected in the simulated antibody profiles across individuals. Grey lines are the distributions of peak frequencies of the 1,000 permutations of the simulated antibody profiles. For each scenario, we simulated the life-course of infections and immune responses for 777 individuals of the same age as the participants in our study and extracted the antibody profile in 2014 for the year’s corresponding to when our twenty strains were isolated. **(A)** No biological mechanisms were modeled, and the individual risk of infection each year was purely random with a fixed probability of 0.2. **(B)** Narrow (i.e., against antigenically similar strains) and broad (i.e., against distant strains) cross-reactions of antibodies were modeled, which would however not affect individual risk of infection every year (i.e., the risk of infection each year was purely random with a fixed probability of 0.2). **(C)** Individual risk of infections was modeled as the randomly varied population-level H3N2 activity every year (i.e., not affected by individual antibody responses), no cross-reactions of antibodies were modeled. **(D-F)** Narrow and broad cross-reactions of antibodies were modeled, with greater cross-reactions conferring higher level of protection. Population-level H3N2 activity were modeled as constant **(D)**, randomly **(E)** and periodically **(F)** varied, respectively. **(G)** Broad cross-reactions of antibodies were modeled, with greater cross-reactions conferring higher level of protection. Random variations in population-level H3N2 activity were modeled. **(H)** Narrow cross-reactions of antibodies were modeled, with greater cross-reactions conferring higher level of protection. Random variations in population-level H3N2 activity were modeled. **(I)** Biological mechanisms included in models that generated results in panel A to H.

We simulated individual infection histories since 1968 and sampled these simulated histories with the same time resolution as tested strains measured in 2014. We applied Fourier analysis on the resulting individual time series (see Methods). We tested several potential biological mechanisms that can shape individual antibody profiles through influencing individual infection hazard (i.e., individual pre-existing titer to the circulating strain and population-level circulation) and antibody responses after exposures (i.e., broad, and narrow cross-reactions) (Fig. 3I and Fig. S13). The breadth of such cross-reactions was implicitly assumed to be determined by the antigenic evolution rate in our simulations, which is 0.778-unit changes in the antigenic space per year according to prior estimates (Fonville et al., 2014; Kucharski et al., 2018).

We assessed the periodic pattern of the simulation from two perspectives. First, we compared whether the peak frequency distribution from the simulation was significantly different from the null distribution, to determine if the simulated antibody profiles were periodic (Figure 3I). Next, we compared whether the simulated antibody responses had a higher proportion of peak frequency of 0.025 to 0.050 compared with the null distribution, to determine if the simulations could recover the long-term periodicity that was identified in the empirical data.

Multiple models showed qualitatively similar periodic behavior to data, that is different from null distribution and had significantly higher proportion of simulated individual responses with long-term periodicity (Fig. 3). A key model component that exhibited long term periodicity was cross-reactivity between antigenically similar viruses, especially broad range (i.e., against distantly related strains) cross-reactions (Fig. 3D-H). When the component of broad range cross-reactions was absent in the model, population-level circulation alone was not able to recover the long-term periodicity in individual antibody responses (Fig. 3B). However, when cross-reaction in antibody responses was included in the model, a less predictable population-level activity (i.e., random compared to cyclic variation, Fig. 3G-H) appeared to introduce more uncertainties in the observed cycles in individual antibody responses.

### Predicting seroconversion to recent strains using cycles in individual antibody responses

These results suggested that, after accounting for the impact of population-level A(H3N2) circulation, cross-reactivity from previously infected strains likely explained the reported cyclic patterns in an individual’s antibody responses. As such, we hypothesized 1) that the position of individuals in their antibody response cycles could be predicted years in advance if the periodic behavior was stable over three-four years and 2) that the position of individuals in their antibody response cycles are associated with responses to future strains. We measured the position in antibody response cycles using phase angles (Fig. 4A)

**Figure 4.**
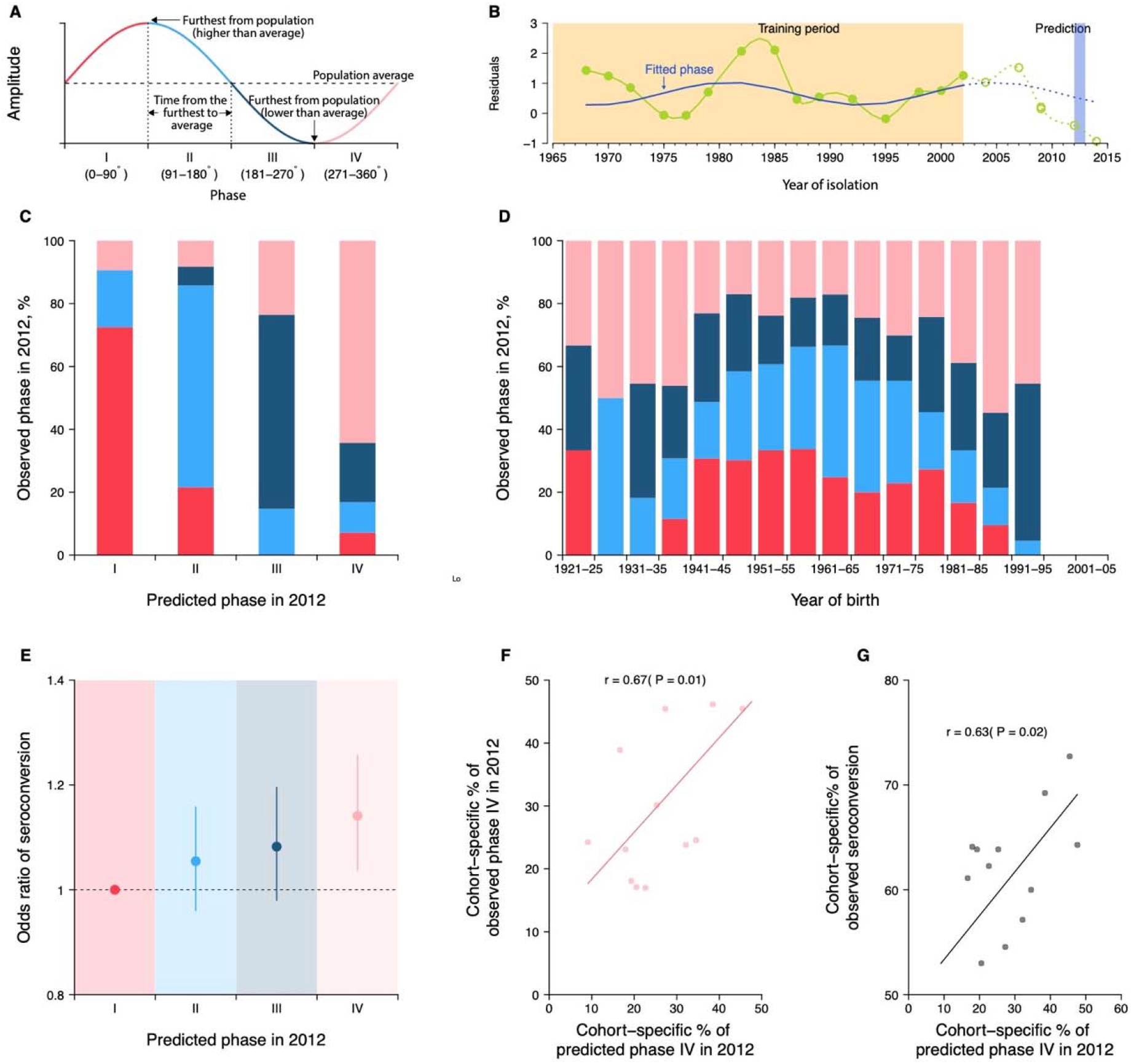
Predicting seroconversion to the recently circulated strains using phases of individual antibody responses. **(A)** Concept plot for phases. Four phases were classified based on the phase angles between 0 to 360 degrees. **(B)** Illustration of predicting phase in 2012 using individual residuals from baseline HI titer that were measured against 14 historical strains (i.e., isolation year up to 2002). Green dots and solid green lines indicate the residuals against historical strains that were used to fit the periodic function (shown in solid blue lines). With the fitted periodic function, we predicted phase angles in 2012 based on the predicted residuals for individual’s titers against strains that were circulating after the training period (shown in dotted blue lines). For reference, we also showed the observed baseline residuals for individual’s titers against strains that were circulating after the training period (indicated as green circles and dotted green lines). **(C)** Observed and predicted phase in 2012 across participants. Colors represent participants’ observed phase in 2012, with I, II, III and IV represented by red, light blue, dark blue and pink, respectively (same for D and E). **(D)** Observed cohort-specific distribution of phase in 2012. **(E)** Adjusted risk of seroconversion to recent strains (i.e., A/Texas 2012 or A/HongKong/2014) between baseline and follow up visits for different phases. We estimated associations between phases in 2012 that were estimated from individual antibody profile residuals and seroconversion to any of the two recent strains and adjusted for age at sampling and the average pre-existing titer of the two strains. **(F)** Observed and predicted cohort-specific proportion of phase IV in 2012. **(G)** Predicted proportion of phase IV in 2012 and the observed proportion of seroconversion to recent strains between baseline and follow up for each cohort.

To test the first hypothesis, we predicted the phase of individual antibody response residuals to strains circulating in 2012 (mid-point of baseline and follow-up; Fig. 4B), by fitting a log-linear regression to the residuals of HI titers measured at baseline (2010) against 14 historical strains (i.e., isolated between 1968 and 2002) on harmonic terms that represent the long-term periodicity (assuming as 24 years). We found high consistency between predicted and observed phases in 2012 across participants (Fig. 4C). For example, a consistency of 73% (95% CI, 65% to 79%) among individuals whose antibody responses were predicted to be in phase I (i.e., the first quarter of a cycle).

To test the second hypothesis, we fitted a logistic regression of seroconversion (i.e., four-fold rise in HI titers to A/Texas/2012 or A/HongKong/2014) between baseline and follow-up on the above-mentioned predicted phase in 2012 (using strains isolated between 1968 and 2002), and adjusted for biological age at baseline and the average pre-existing log-titer of the two strains. We found that individuals who were predicted to be in phase IV (i.e., the last quarter of a cycle) were 14% (95%CI, 4% to 26%) more likely to experience seroconversion to the two recent strains, compared to those in phase I (Fig. 4D).

### Disentangling cohort effects using cycles in individual antibody responses

As results of resonance, we expected intrinsic cycles in individual antibody responses to be correlated across birth cohorts (Fig. 4E). There were indications of this in the correlation of phase across cohorts. For example, we observed a higher proportion of participants who were in phase IV for the 2012 strain at baseline, when comparing the birth cohorts of 1986-90 (55%, 95% CI, 39% to 70%) with 1961-65 (17%, 95% CI, 11% to 25%; chi-squared test, P < 0.001). Moreover, we found that such cohort-specific differences in phase composition (Pearson correlation = 0.67, P = 0.01; Fig. 4F) and the resulting proportions of seroconversion (Pearson correlation = 0.63, P = 0.02; Fig. 4G) correlated with the predicted cohort-specific composition of phase IV in 2012. However, we also found that a diversity of phases was exhibited by members of the same age cohorts, suggesting that cycles of individuals could depart from members of their own birth cohort.

## Discussion

We demonstrate that human antibody responses to influenza A(H3N2) display long-term periodicity, which are biologically consistent with non-linear human adaptive immune responses (i.e., cross-reactions) to evolving viruses. Our observations are validated by different analytic methods and validation in a separate study population tested by a different antibody assay. Our findings were robust to our sampling and interpolation methods (Fig. S9 and S10). We further demonstrate that, at both individual and birth-cohort levels, the phases of the antibody responses to the currently circulating strains are predictable and associated with seroconversion to these strains independent from the pre-existing titers and age. Such findings could improve our forecasting of the individual and birth-cohort level risks of infections and our understanding of heterogenous immune responses and vaccine effectiveness against influenza viruses.

We were able to qualitatively recover the observed long-term periodicity only when including cross-reactions between antigenically similar strains in the simulations. Such findings fit in previous observations that strain-transcending antibody responses to past infections accumulate and build up contemporary antibody profiles (Fonville et al., 2014; Yang et al., 2020). Within-subtype cross-reactions may drive individual-level long-term periodicity in antibody responses through temporal (but waning) cross-protection (i.e., positive feedback) and blunting generation of specific antibodies (i.e., negative feedback) against the circulating strains. A recent cohort study found homotypic cross-protection against PCR-confirmed infections for up to five seasons after infections supported cross protection that eventually wanes (Wraith et al., 2022). In addition, the approximately 24-year periodicity implicitly suggested that antibodies gained from last immunizing events may interfere with the antibodies against the circulating strains for a maximum 18 years (i.e., phase I to III in Fig. 4B), before the antigenicity between the last immune strain and the circulating strain was too different to cause high-level cross-reactions in binding antibodies. This is in line with our previous findings that people’s sera showed very little to no cross-reaction with strains that were isolated 20 years prior to their births (Lessler et al., 2012; Yang et al., 2020).

Our findings suggest that long-term periodicity in HI titer may be driven by broad cross-reactions between strains that accumulate as people are exposed to multiple viruses over their lives. The breadth of such cross-reactions was determined by previously reported antigenic evolution (Fonville et al., 2014; Kucharski et al., 2018). In simulations, we found that antigenic evolution rates significantly change the periodicity in individual antibody responses (Fig. S14). Slower antigenic evolution rates shift cycles in individual antibody responses to longer periodicity (Fig. S14), with the extreme that people could acquire life-long immunity against antigenically stable viruses (e.g., measles) (Amanna et al., 2007). Though in our simulations, faster antigenic evolution led to shorter cycles, high rates of antigenic evolution could diminish the periodicity in antibody responses through frequent reactions to re-exposures (Fig. S14).

We found associations between the phase of antibody response cycles and the risk of seroconversion to circulating strains, after accounting for the homologous pre-existing HI titers. Previous studies have reported differential risk in individuals with the same homologous HI titer, proposing that unknown individual exposure histories and cohort-effects as possible explanations (Turbelin et al., 2013; Yang et al., 2018). Our findings suggest that cyclic patterns in an individual’s antibody responses, which may be predictable at both individual and cohort level, may contribute to this heterogeneity. In addition, our findings suggested that measuring seroconversion against a circulating strain could reveal a limited amount about protective immunity. Measuring responses to both circulating and previously circulating viruses (as well as calculating phase) could improve characterization of people’s risk (Quandelacy et al., 2021).

We demonstrate that resonance of cycles in individual-level antibody responses could form variations in phase distribution of antibody responses across birth-cohorts, which is consistent with previous findings that the fraction of A(H3N2) associated cases across different birth cohorts was found to change year to year (Turbelin et al., 2013; Yang et al., 2018). Our results also showed that the phase distribution of antibody responses across birth-cohorts may be predictable and could be further used to predict the cohort-specific seroconversion against the circulating strains. This is potentially useful to determine and vaccinate the high-risk groups based on the exposure histories that could be shared by birth-cohort. We did not attempt to explore the impact of antigenic evolution speed and the associated dynamics in antibody responses on shaping age-specific patterns of cases, while we speculate that antigens with faster antigenic evolutions may attack different age groups at relatively similar risks, while antigens with slower antigenic evolutions tended to attack the children (e.g., A(H3N2) vs. B/Victoria in (Turbelin et al., 2013; Yang et al., 2018)).

We recognize that HA-binding antibodies only mediate about half of the protection against influenza infections, while other forms of immunity, including neutralizing antibodies, non-HA head specific inhibitory antibodies and cellular immunity, would provide independent protections against infection and the severity of the diseases (Cowling et al., 2019; Meade et al., 2020; Ng et al., 2019; Stadlbauer et al., 2019). The breadths and oscillation patterns may differ across different forms of immunity.

Our work has several limitations. First, between-subtype interactions have not been incorporated into our framework. It is arguable that whether infections with A(H1N1) could confer years-long cross-protections against A(H3N2) in human at individual level (Sonoguchi et al., 1985), while prior findings tended to support that between-subtype interaction could alter the transient cycles (i.e., within seasons) (Goldstein et al., 2011; Meade et al., 2020; Ranjeva et al., 2019). Nevertheless, our simulation results suggested that only including population-level circulation – regardless of its underlying drivers - could not recover the observed long-term periodicities in individual antibody responses. Second, our simulation results, while robust across parameter settings, may depend on the simplifying assumptions on immunological mechanisms we made (e.g., individual immunity-dependent protection) and therefore only qualitatively recovered the observed pattern. Finally, the exact value for the long-term periodicity was determined by a series of fixed frequencies that were examined in the Fourier analysis, which depend on the number and span of the tested strains. Therefore, more accurate values for the periodicity could be estimated if the tested strains were sampled more densely.

## Materials and methods

### Cohort and serological data

We used serum collected from 777 participants who were recruited to an ongoing Fluscape cohort in Guangzhou, China, and provided blood samples for both a baseline visit (December 2009 to January 2011) and a follow-up visit (June 2014 to June 2015) (Jiang et al., 2017; Yang et al., 2020). The cohort recruited 40 locations that are randomly distributed in a fan-shape area spanning from the city center to the neighboring rural areas. Participants, aged 2 to 86 years old at baseline sampling, were recruited from households that were randomly selected in these locations. Details of the cohort and participants included have been described previously (Jiang et al., 2017; Yang et al., 2020).

We measured antibody titers against 21 A(H3N2) strains using hemagglutination inhibition (HI) assays of paired serum collected from the two visits (Yang et al., 2020). Strains tested were isolated from 1968 to 2014 (Yang et al., 2020) and priority was given to those included in vaccine formulation and/or used to construct the antibody landscape by Fonville et al. (Fonville et al., 2014). The strains we used are A/Hong Kong/1968, X-31 (isolated in 1970), A/England/1972, A/Victoria/1975, A/Texas/1977, A/Bangkok/1979, A/Philippines/1982, A/Mississippi/1985,A/Sichuan/1987, A/Beijing/1989, A/Beijing/1992, A/Wuhan/1995,A/Victoria/1998, A/Fujian/2000, A/Fujian/2002, A/California/2004, A/Brisbane/2007, A/Perth/2009, A/Victoria/2009, A/Texas/2012, A/Hong Kong/2014. Detailed laboratory methods have been described previously (Lessler et al., 2011; Yang et al., 2020).

### Statistical analysis

#### Generalized additive model

To extract population-from individual-level A(H3N2) activity, we fitted a generalized additive model (GAM) of log HI titers (Fig. 1A and Fig. S1A) on the spline of age at baseline sampling, the spline of age at circulation (i.e., difference between year of strain isolation and year of birth of the participant) with strain-specific intercepts, which has been described in detail in a previous study (Lessler et al., 2012). In brief, log titer for strain *j* and participant *i* is modeled as:

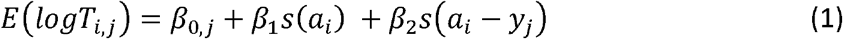

where *s*(.) denotes spline terms. *a*_*i*_ denotes the age of the participant *i* at baseline sampling and *y*_*j*_ denotes the number of years since strain *j* was isolated until baseline sampling. Strain-specific intercepts *β*_0,*j*_ were estimated and further used as a proxy for the population level variations in A(H3N2) activities between 1968 and 2014 in the main analysis (Fig. 1B and Fig. S1B).

Residuals were calculated as the difference between observed and predicted log titers from the fitted GAM to characterize individual level A(H3N2) immune responses (Fig. 1C and Fig. S3A). A time series of residuals for participant *i* was derived as:

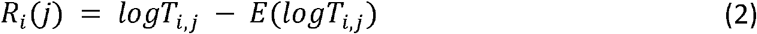

and then chronologically ordered by the year of strain isolations. HI titers for baseline and follow-up visits were fitted separately, and only titers to strains that were isolated after the person was born were included in the model.

#### Fourier analysis

Periodicity in individual antibody responses to influenza was examined using Fourier spectral analysis with linear detrending, from which variances explained by each frequency were extracted (Fig. 1D and Fig. S3B). As the tested A(H3N2) strains were irregularly spaced in time (two to three-year intervals), we fitted a spline and interpolated the time series to a yearly resolution before applying the Fourier analysis.

For individual level periodicity, we extracted the frequency that explained the most variance (i.e., the greatest spectral power; ‘peak frequency’ hereafter) for each individual (Fig. 1D and Fig. S3B) and plotted the distribution of peak frequencies across 777 participants (Fig. 1E and Fig. S1C). To test the significance against the null distributions, we compared the observed distribution of peak frequencies with the distribution of peak frequencies from 1,000 permutations. In each permutation, we shuffled the time series of residuals for each person and extracted the peak frequency for each individual (Fig. S3C-D).

### Validations and sensitivity analyses

#### Weighted frequency

The peak frequency of the Fourier spectrum we extracted only represents the frequency that explained the most variance, but it cannot reflect the variance explained by the other frequencies, i.e., whether the Fourier spectrum is skewed towards the peak frequency or is flatly distributed (Fig. 1D and Fig. S3). Thus, we calculated the average frequency weighted by the variance explained (“weighted frequency”), to represent the weighted center for each spectrum (Fig. S3B). The weighted frequency *f*_*w*_ was calculated as:

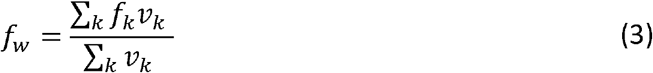

where *f*_*k*_ and *v*_*k*_ denote the *k*^*th*^ examined frequency and its estimated variance the Fourier spectrum. We found our data was more likely to show lower weighted frequencies and longer periods compared to the permutations (Fig. 2A).

#### Addressing irregularly sampled intervals with Lomb-Scargle periodogram

To examine the impact of using irregularly sampled intervals and interpolation on the results from the Fourier spectrum analysis, we performed a sensitivity analysis using the Lomb-Scargle periodogram (Glynn et al., 2006), which is often used to detect the periodicity of irregularly sampled time series. As in the previously described Fourier spectrum analysis, we derived the spectrum for each individual’s time series of residuals using the Lomb-Scargle periodogram, which estimated the variance explained at each frequency. We then extracted the frequency with the most variance explained, i.e., “peak frequency” for each spectrum. Similar to the main analysis, we compared the distribution of observed peak frequency derived from Lomb-Scargle periodogram across participants with those from 1,000 permutations (Fig. 2B). In each permutation, we shuffled the time series of residuals for each individual (maintaining the irregularity in the sampling), and then extracted the peak frequency of the Lomb-Scargle periodogram for each shuffled time series.

#### Removing non-linear trends with Empirical Mode Decomposition (EMD)

Although we removed the linear trend before applying Fourier analysis, several time series contained non-linear trends that could potentially bias the estimate of the peak frequency to lower values (e.g., participant 1 and 2 in Fig. S8). In order to avoid this issue, we performed Fourier analysis with the time series of residuals after removing non-linear trends using Empirical Mode Decomposition (EMD)(Huang et al., 1998).

To do this, we first applied EMD to each individual’s time series of residuals, and extracted the underlying trend, defined as the “residue” remaining after all intrinsic mode functions have been extracted (Fig. S8). We then detrended the time series by subtracting this “residue” from the original time series (Fig. S8). Finally, the peak frequency of the Fourier spectrum of the detrended time series was extracted for the individual, and the distribution of peak frequencies was plotted across individuals (Fig. 2C). For each permutation, we shuffled the individual’s time series and applied EMD to the shuffled time series. The remaining steps for the permutation analysis were the same as above.

For participants whose time series showed non-linear trends (e.g., participant 1 and 2 in Fig. S6), peak frequency shifted to a higher frequency after detrending with EMD. Meanwhile, for participants whose time series showed cycles (e.g., participant 3 and 4 in Fig. S8), the low frequency cycles were no longer detectable after detrending with EMD. Therefore, the results shown in Fig. 2B were the distribution of peak frequencies after removing both non-linear trends and some low frequency cycles. The 20-40 years cycles were still detectable for both visits, suggesting the long-term cycle we detected was not solely explained by the non-linear trend of the time series.

#### Dropping every other strain

In order to test whether the reported cycles in the individual residuals were influenced by the relatively stronger responses to some strain (e.g., X-31, A/Mississippi/1985, A/Beijing/1992 and A/Fujian/2002), we dropped one out of the twenty-one strains and repeated the Fourier analysis to the time series of the remaining twenty strains. For the permutation test, we shuffled the time series of the remaining twenty strains and re-interpolated the shuffled time series for each individual. Results suggested that dropping out one strain did not affect our conclusions (Fig. S9 and S10).

#### Validations using random values or values from periodic curves

We tested the robustness of our results from the Fourier analysis with a time series of twenty-one irregularly sampled data points with the same time resolution as our data. Time series consisted of random values generated from varying underlying distributions. Briefly, we drew a set of random values for each individual and the length of time series was based on the individual’s year of birth. We performed the interpolations, Fourier analysis and extracted the peak frequency of the Fourier spectrum for each new time series. Finally, the distribution of peak frequencies for the simulated time series and their null distributions from permutations were compared.

We performed this analysis using values drawn from normal and lognormal distributions without periodicity (Fig. S6A-B). In addition, we randomly replaced 2 to 4 points in each individual’s time series with outlier values that are rare in the underlying distribution, in order to mimic the relatively higher titers to several strains observed in the data (Fig. S6C-D). There were no significant differences between peak distributions of the simulated random time series and their permutations, suggesting the low frequencies identified in our real data cannot be explained by the correlation structure introduced by irregularly sampled intervals, interpolation, randomness, and outlier values.

We then applied the Fourier analysis on time series generated by sampling from periodic curves with white noise. To do this, we first simulated a time series from 1968 to 2014 on a yearly basis for each participant from a sinusoidal curve with a certain periodicity and white noise. We then subset the simulated time series to the years when our tested A(H3N2) strains were isolated relative to each participant’s year of birth. We applied the previously described interpolation and Fourier analysis to the subset of each time series. We repeated the above analysis for 777 participants and compared the distributions of peak frequencies from simulated time series and their 1,000 permutations. Four scenarios were tested: 1) time series of all participants had a single 25-year periodicity (Fig. S7A); 2) time series of all participants had a single 16-year periodicity (Fig. S7B); 3) time series of half of the participants had a single 25-year periodicity, and time series of the other half of participants had a single 16-year periodicity (Fig. S7C); and 4) time series of all participants contained two superimposed periodic curves, with periodicities of 25 and 5 years (Fig. S7D). Results (Fig. S7) suggested that the method we used in the main analysis can uncover the real low frequency signals, while uncovering high frequency signals could be challenging due to the resolution of our data.

#### Excluding participants who were born after 1968

To examine the effects of participants who had a relatively shorter exposure history of A(H3N2) on the reported cycles, we repeated the Fourier spectrum analysis with time series of residuals for a subset of participants (n = 487) who could have experienced all tested A(H3N2) strains, i.e., born before 1968. The analysis follows the same steps as the main analysis except that the distributions of peak frequencies were plotted across 487 eligible participants. Cycles with low frequencies were found for the subset of senior participants as well, with an increasing proportion of participants having the lowest frequency (Fig. S11).

#### Sera from Vietnam study

In order to test our results with a different population, we repeated the analysis with publicly available data reported in a previous Vietnam study (*4, 21*). Longitudinal sera were collected for 69 participants in Ha Nam, Vietnam. Participants were aged 7 to 95 years old in 2012, of which 48% were under 30s (Fonville et al., 2014). Sera were repeatedly collected from these participants between 2007 and 2012 on a yearly basis (Fonville et al., 2014). HI titers were measured for 57 A(H3N2) strains isolated between 1968 and 2011, with a finer resolution in the more recent years (Fonville et al., 2014).

In the Vietnam study, multiple strains had been isolated in the same year, resulting in multiple titers being available for a given year for each individual. Therefore, we fitted a cubic spline in order to derive a time series that captured the geometric mean titers to strains isolated in the same year. We then applied Fourier analysis to each splined time series and extracted the peak frequency of each spectrum. The distribution of peak frequencies was characterized across 69 individuals by the year of serum collection (Fig. S12). For the permutation analysis, HI titers were shuffled before fitting splines to the time series. As the age of participants was not available, we performed the analysis with raw titers without adjustment on age. Significant cycles with frequencies ranging from 0.050 to 0.075 (∼ 13 to 20 years) were detected for serums collected in 2007, 2009, 2010 and 2011, coinciding with the frequencies detected using the raw titers of serums collected in our baseline visit.

### Simulations of life-course infection history and immune responses

#### Model descriptions

In order to explore the mechanisms behind the reported dynamics of human immune responses to influenza, we applied a previously described mechanistic model (Kucharski et al., 2018) to generate realizations of lifelong infection history and subsequent immune responses. Simulations were individual based on a yearly scale and returned as antibody profiles consisting of titers to a panel of 47 strains (i.e., strains isolated from 1968 to 2014) that were tested in 2014. The simulations consisted of the following steps:

1. *Construct initial antibody profile*. An initial antibody profile was generated for the sera collected in 1968 for participants who were born on or before 1968, or the year of birth for participants who were born after 1968. Titers to all 47 strains were assumed to be 0 for initial antibody profiles.
2. *Extract pre-existing titers for each season*. For an examined year *y*, we extracted the titer to the strain that was isolated in *y* from the latest antibody profile (i.e., antibody profile measured in year *y*) (Fig. S13).
3. *Determine the probability of infection of the circulating strain*. The probability of an individual infected by the strain isolated in year *y* was calculated according to the immunity-dependent protection (Eq. 11; see section “Modeling immunity-dependent protection” for details) and annual A(H3N2) activity (Table S2). For the initial year (i.e., 1968), we imposed a pandemic with an attack rate of 50% in the main analysis. The strain isolated in year *y* was assumed to be the circulating strain of that year.
4. *Simulate infection event*. Infection outcome was randomly generated following a binomial distribution with the probability calculated in step 3). Infection outcomes were simulated for each individual every year.
5. *Update immune responses*. Immune responses (i.e., boost and cross-reactions from infections and/or immunity decay) to the whole panel of strains were updated based on the annual infection outcome (Fig. S13) using the previously described model and estimates (Eqs. 4-9 and Table S2; see section “Modeling immune responses” for details)(Kucharski et al., 2018). The updated antibody profiles are then used in step 2) for the following year (Fig. S13).
6. Repeat steps 2) to 5) until 2014 and extract the antibody profiles measured in 2014.

We simulated antibody profiles for 777 individuals of the same ages as the participants in our study. For each individual, we repeated the above 6 steps from 1968 (or the year of birth) to 2014 and extracted the antibody profiles to all 47 strains in 2014 for further analyses.

In order to explore the mechanisms that created the observed cycles, we performed simulations under different scenarios that considered several generally recognized components of immunity (Fig. 3I):

1. Baseline scenario (Fig. 3A), which assumed a constant 50% annual probability of infection for all individuals and no cross-reaction or cross-protection from past infections.
2. Population activity only scenario (Fig. 3B), which assumed a random varied population-level viral activity that would affect individual probability of infection, and no cross-reaction or cross-protection from past infections.
3. Narrow cross-reaction scenario (Fig. 3C), which assumed annual individual probability of infection would be determined by individual pre-existing titer and a random varied population-level viral activity, and cross-reactions only to a narrow range of antigenic relatives (i.e., recent strains).
4. Broad cross-reaction scenario (Fig. 3D), which assumed annual individual probability of infection would be determined by individual pre-existing titer and a random varied population-level viral activity, and cross-reactions only to a broad range of antigenic relatives (i.e., distant strains).
5. Cross-reaction only scenario (Fig. 3E), which assumed a constant 50% annual probability of infection for all individuals, and cross-reaction to both narrow and broad range of antigenic relatives, but no cross-protection from past infections.
6. No population activity only scenario (Fig. 3F), which assumed annual individual probability of infection would be determined by individual pre-existing titer but not population-level viral activity, and cross-reactions and cross-protection to both narrow and broad range of antigenic relatives.
7. Random population activity scenario (Fig. 3F), which assumed annual individual probability of infection would be determined by individual pre-existing titer and a random varied population-level viral activity, and cross-reactions and cross-protection to both narrow and broad range of antigenic relatives.
8. Periodic population activity scenario (Fig. 3F), which assumed annual individual probability of infection would be determined by individual pre-existing titer and a periodically varied population-level viral activity (5-year periodicity), and cross-reactions and cross-protection to both narrow and broad range of antigenic relatives.

#### Modeling immune responses

We adapted the previously described model to simulate immune responses after exposures(Kucharski et al., 2018); the parameters used are shown in Table S2. The immune response after an infection is divided into long-term boosting, *d*_*l*_ (*j, m*_*t*_) and short-term boosting, *d*_*s*_ (*j, m*_*t*_), modeled as:

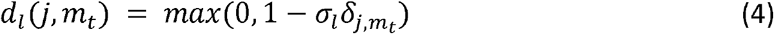

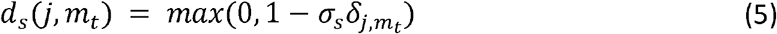

where 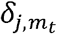 denotes the difference in antigenic difference between strain *j* and the previously infecting strain *m*_*t*_:

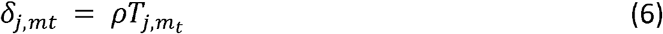

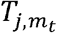 denotes the number of years between when the tested strain *j* and the infected strain *m*_*t*_ were isolated, and *ρ* is the rate of change in antigenic units per year. Parameters *σ*_*l*_ and *σ*_*s*_ represent the durations of cross-reactions. Short-term immunity also wanes, as set by the waning duration *ω* and the number of years between the year of infection by strain *m*_*t*_ and year of testing 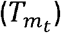:

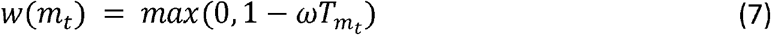

The antigenic seniority was scaled by a suppression parameter *τ* and the order of infection (*N*_*m*_) among all infected strains *X*_*t*_:

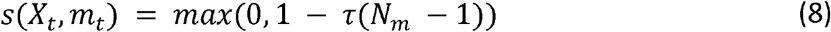

Prior study estimated *τ* as 0.04, while we explored both 0 and 0.04 and found minimal impact on our main results. Therefore, we assumed *τ* as 0 for simplicity.

Finally, the titer against strain *j* for person *i* tested in year *t* is:

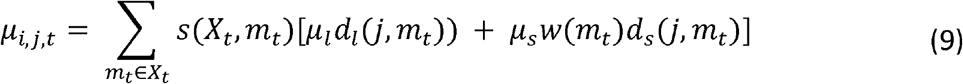

where *μ*_*l*_ and *μ*_*s*_ denote the mean log titers of long-term and short-term boost to an infecting strain, respectively.

#### Modeling immunity-dependent protection

For the baseline scenario and cross-reaction only scenario, the probability of infection was assumed to be a constant. For the cross-protection and antigenic seniority scenarios, a higher HI titer to a circulating A(H3N2) strain is assumed to be associated with lower risk of infection with that strain (Fig. S13). We assumed that the 50% protective titer is 1:40 (i.e., *μ*_50_ = 3 on a log scale). The titer-dependent risk of infection is modeled as (*37*):

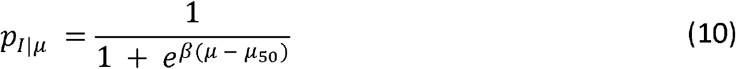

where *β* is the scale parameter of the titer-dependent protection estimated in previous studies (Yuan et al., 2017). After adjusting for annual A(H3N2) activity (*λ*_*t*_), the titer-dependent probability of infection of strain *j* for person *i* tested in year *t* is:

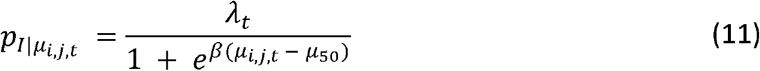

The annual A(H3N2) activity, *λ*_*t*_, was included to explore the impact of the virus circulation at population level on the observed long-term cycles in individual antibody responses. Three different hypothetical scenarios were assumed for *λ*_*t*_:

1. *λ*_*t*_ = 2, where annual activity was assumed as constant across the 47 years with an annual attack rate of 20%.
2. *λ*_*t*_ ∼ *Uniform*(0, 0.2), where annual activity varies between 0 and 0.2 randomly.
3. *λ*_*t*_ = 0.2*sin* (2*πt*/5), where annual activity varies year to year with a periodic pattern.

The main objective of this analysis was to demonstrate that population circulation alone was not able to recover the observed periodicity in individual antibody responses. As thus, although there remain debates about the interactions between influenza subtypes, we showed that it seemed not to be the main driver of the observed periodicity in individual antibody responses.

### Prediction of individual antibody responses to future strains using intrinsic cycles

#### Estimation of the phase

We estimated the phase angle (*p*_*y,i*_, in degree) of antibodies against a strain *j* that circulated in a given year *y* for person *i*, to represent the position where the antibody against the tested strain stands in the entire antibody responses of this person. We first fitted a regression to the time series of individual residuals (*R*_*i*_(*y*)) for strains that were isolated during a certain period and included harmonic terms that represent the periodic patterns in the antibody responses.

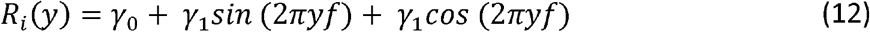

where *f* is assumed as the inverse of the periodicity that most of our participants showed, i.e., 24 years. With the estimated coefficients from Eq. 12, we predict the phase angle in radian (*r*_*y,i*_) of strain *j* that circulated in given year *y* for person *i* as follows:

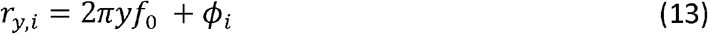

where *ϕ*_*i*_ denotes the person *i* ‘s phase shift:

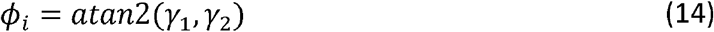

We then translated the phase angle from radian to degree as follows:

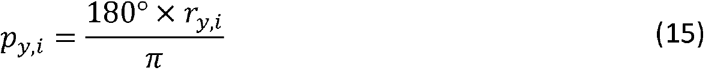

The phase angle in degree was then classified into four categories, namely, phase I (0-90°), II (91-180°), III (181-270°) and IV (271-360°) (Fig. 4A).

#### Comparison between observed and predicted phase in 2012

We predicted the phase angle (in degree) for the strain that circulated in 2012, which is the middle between our baseline (2010) and follow up (2014) visit. We first fitted Eq. 12 to HI titers that were measured for strains isolated between 1968 and 2002 measured at baseline (i.e., 14 strains, Fig. 4B). Predicted phase in 2012 was then estimated using Eqs. 13 to 15. To estimate the observed phase in 2012, we fitted the model in Eq. 12 to the full panel of tested strains (i.e., 21 strains) measured at baseline, and calculated the phase angle using Eqs. 13 to 15.

To assess the consistency between the prediction and observation of phases in 2012, we plotted the distribution of the observed phase in 2012 among people who were predicted to in each of the four phases in 2012 (Fig. 4C).

#### Association between phase and seroconversion

We examined the association between the phase in individual antibody responses and antibody responses to circulating strains (Fig. 4D). We measured the antibody responses circulating strains as the seroconversion (i.e., fold of change ≥ 4) to either A/Texas/2012 or A/HongKong/2014 (i.e., strains that were circulated between baseline and follow up). We fitted a logistic regression to seroconversion and adjusted for the predicted phase (in categories) in 2012, the average of titers against the two tested strains at baseline (i.e., pre-existing titers in log scale) and the participants’ age at baseline.

### Disentangle birth cohort effects using intrinsic cycles in individual antibody responses

To examine the differences in phase distribution across different birth cohorts, we first estimated the observed individual phase in 2012 by fitting Eqs. 12-15 to the full panel of tested strains measured at baseline. We compared the distribution of phase in 2012 among 5-year binned birth cohorts using chi-squared test (Fig. 4E).

We estimated the phase distribution across birth cohorts using the predicted phase in 2012, which was derived by fitting Eqs. 12-15 to the HI titers against strains isolated between 1968 to 2002 measured at baseline. We examined the association between the predicted and observed phase distribution across birth cohorts by calculating the Pearson correlation between the predicted and observed proportion of phase IV in each birth cohort (Fig. 4F).

We examined the association between the predicted cohort-specific proportion of phase IV and the observed proportion of seroconversion to either A/Texas/2012 or A/HongKong/2014 using Pearson correlation (Fig. 4G).

### Software and programs

All analyses were performed in R Version 4.1.0 (R Foundation for Statistical Computing, Vienna, Austria). We used the ‘mgcv’ package to fit GAMs (Wood, 2011). The Lomb-Scargle periodogram was performed with the ‘spectral’ package (Seilmayer, 2016). We performed the empirical mode decomposition with the ‘EMD’ package (Kim and Oh, 2009). Simulations of life-course infection history and immune responses were performed with the ‘Rcpp’ package (Eddelbuettel and François, 2011).

## Data Availability

All data produced in the present study are available upon reasonable request to the authors.

## Author contributions

Conceptualization: J.L., C.Q.J., J.M.R., K.O.K, Y.G., S.R.; Data curation: B.Y., J.L., R.S.; Formal Analysis: B.Y., B.G.C, D.A.T.C.; Funding acquisition: D.A.T.C, J.L., J.M.R., C.Q.J., Y.G. and S.R. Investigation: R.S., Z.H., Y.G.; Methodology: B.Y., B.G.C, D.A.T.C.; Supervision: D.A.T.C. Visualization: B.Y., B.G.C, D.A.T.C.; Writing - original draft: B.Y., B.G.C., D.A.T.C.; Writing - review, and editing: B.Y., B.G.C, J.L., J.M.R., H.Z., C.J.E.M., J.A.H., K.O.K., R.S., C.Q.J., Y.G., S.R., D.A.T.C..

## Data and material availability

All relevant data and code to reproduce the study findings are available online.

## SUPPLEMENTARY TABLES

**Table S1.** Variance (%) explained by low frequencies and peak frequencies for Fourier spectra of individual residuals.

|  | All participants | Participants born before<br>1968 |
| --- | --- | --- |
| **Low frequency (0.025 to 0.050)** |  |  |
| Baseline | 22.1 (11.1, 35.4) | 19.8 (10.0, 32.2) |
| Follow-up | 21.9 (10.3, 35.5) | 18.7 (9.1, 32.2) |
| **Peak frequency** |  |  |
| Baseline | 36.4 (29.3, 45.1) | 33.9 (28.0, 42.2) |
| Follow-up | 36.8 (30.1, 45.9) | 34.7 (28.6, 42.2) |

**Table S2.** Parameters used in the simulations.

| Parameters | Definitions | Value | Note |
| --- | --- | --- | --- |
| Individual Immune responses |  |  |  |
| $\mu_l$ | Mean log titers of long-term (i.e., broad range) boosting to an infected strain. | 2.02 | Ref ((4)) |
| $\mu_s$ | Mean log titers of short-term (i.e., narrow range) boosting to an infected strain. | 2.69 | Ref ((4)) |
| $\sigma_l$ | Duration in years to determine the decrease of cross-reactions for long-term (i.e., broad range) boosting. | 1/0.130 | Ref ((4)) |
| $\sigma_s$ | Duration in years to determine the decrease of cross-reactions for short-term (i.e., narrow range) boosting. | 1/0.031 | Ref ((4)) |
| $\omega$ | Duration in years for the decay of short-term (i.e., narrow range) boosting. | 1/0.79 | Ref ((4)) |
| $\tau$ | Parameter to determine the antigenic seniority. | 0 | Assumption |
| $\rho$ | Rate of changes in antigenic distance per year. | 0.778 | Ref ((3)). |
| Immunity-dependent protection |  |  |  |
| $\beta$ | Scale parameter of the titer-dependent protection. | 2.1 | Ref ((37)) |
| $\mu_{50}$ | Log titer with 50% probability of protection from infection. | 3 | Ref ((37)) |
| Population-level circulation |  |  |  |
| $\lambda_t$ | A(H3N2) activity in year t. | a) a constant of 0.2;<br>b) random, Uniform (0, 0.2);<br>c) cyclic with a 5-year periodicity and maximum activity of 0.2 | Assumption; Equation 11 |

**Table S3.** Mechanisms examined in the simulations.

| Mechanism | Definition | Parameters | Note |
| --- | --- | --- | --- |
| **Infection risk determinant** |  |  |  |
| Immunity-dependent protection | Lower risk of infection with higher HI titer to a circulating A(H3N2) strain. | $\lambda_t, \beta, \mu_{50}$ | Equation 10 & 11; Fig. S14 |
| Population-level circulation | Constant, random, or periodic fluctuation in population-level circulation of the viruses. | $\lambda_t$ | Equation 11 |
| **Antibody responses** |  |  |  |
| Broad cross-reaction | Interaction of antibody responses to strains that were isolated distantly. | $\mu_l, \mu_s, \sigma_l, \sigma_s, \rho$ | Equation 4 |
| Narrow cross-reaction | Interaction of antibody responses to strains that are antigenically similar. | $\mu_l, \mu_s, \sigma_l, \sigma_s, \rho$ | Equation 5 |

## SUPPLEMENTARY FIGURES

**Fig. S1.**
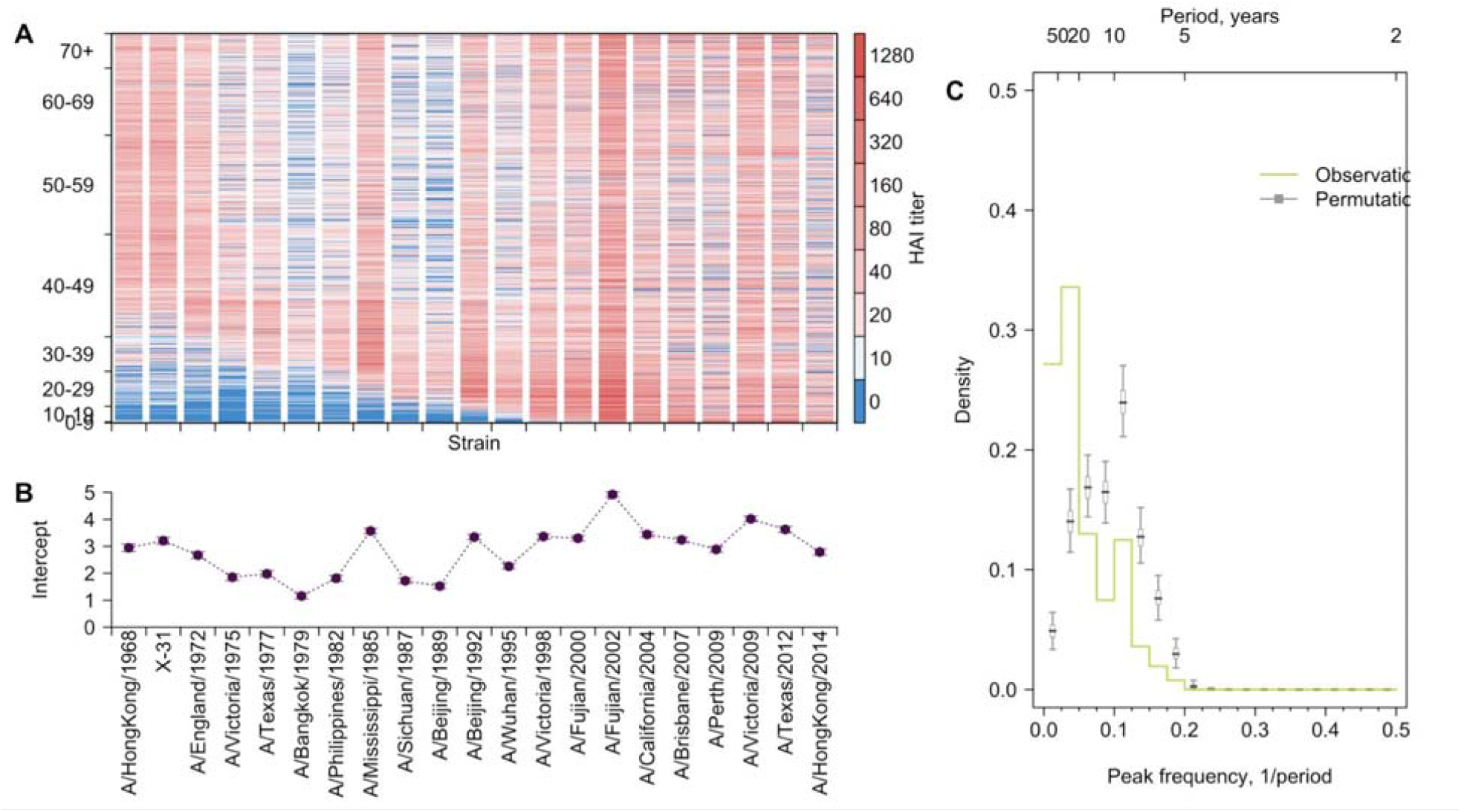
Long-term cycles in individual antibody responses to influenza A(H3N2) at follow-up. **(A)** HI titers against A(H3N2) strains at follow-up. Each row shows an antibody profile for a participant. Participants are sorted by age (y-axis). Strains (x-axis) are sorted by the year of isolation, which are listed in the x-axis of panel B. **(B)** Strain-specific intercepts. A generalized additive model (GAM) was fitted to log HI titers (shown in A) on age at sampling (spline), age at isolation (spline) and strains (categorical). **(C)** Distribution of peak frequencies of individual residuals. We performed Fourier spectral analysis (shown in Fig. 1D) on the time series of residuals of each person and extracted the peak frequency. The light green shows the distribution of peak frequencies across participants. Median (thick grey ticks), interquartile (grey boxes) and 95% intervals (thin grey ticks) of distributions from 1,000 permutations are indicated.

**Fig. S2.**
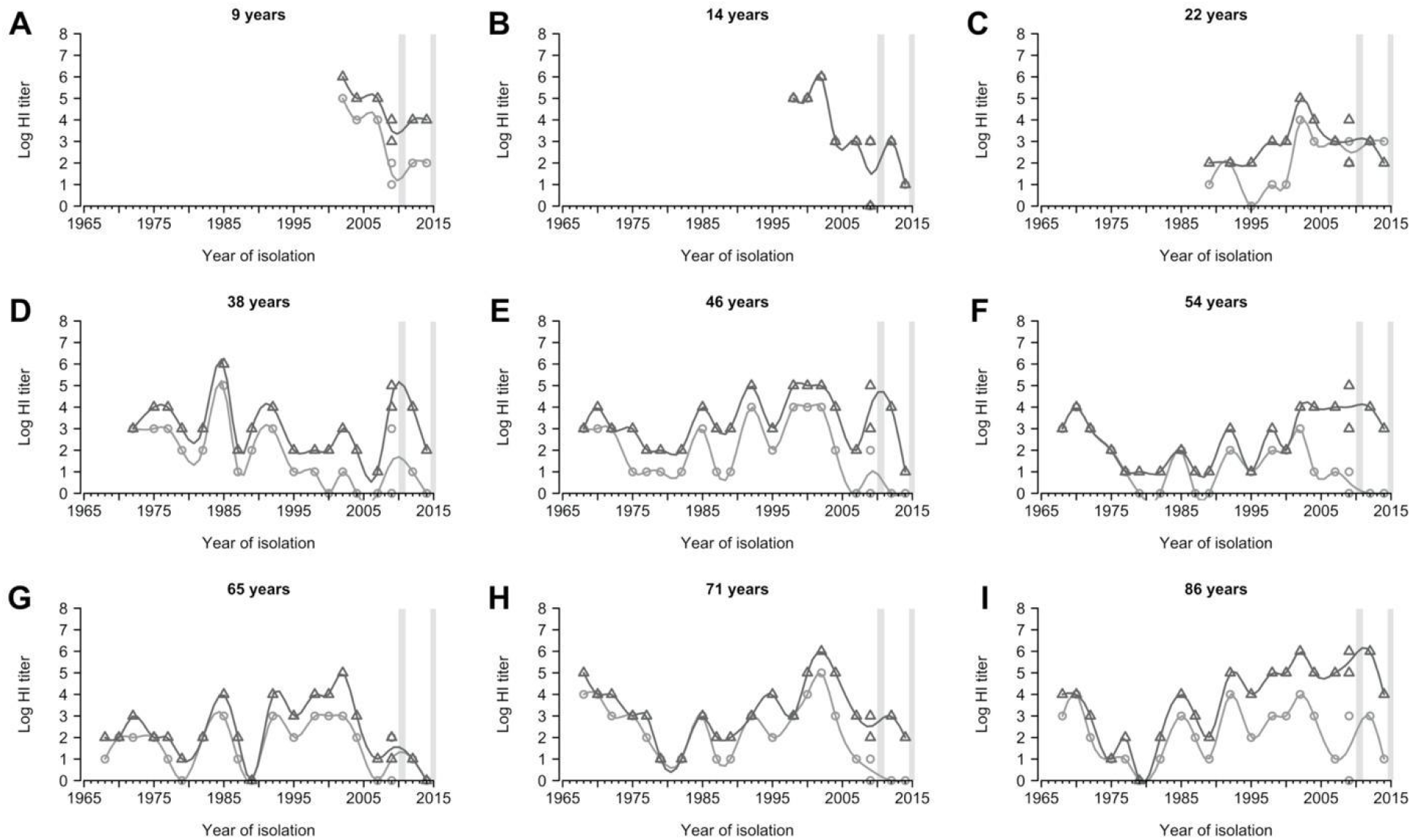
Representative individual profiles of hemagglutination inhibition (HI) titers. Light grey circles and dark grey triangles are antibody measures for baseline and follow-up visit, respectively. Grey vertical boxes indicate the durations of the baseline and follow-up visits.

**Fig. S3.**
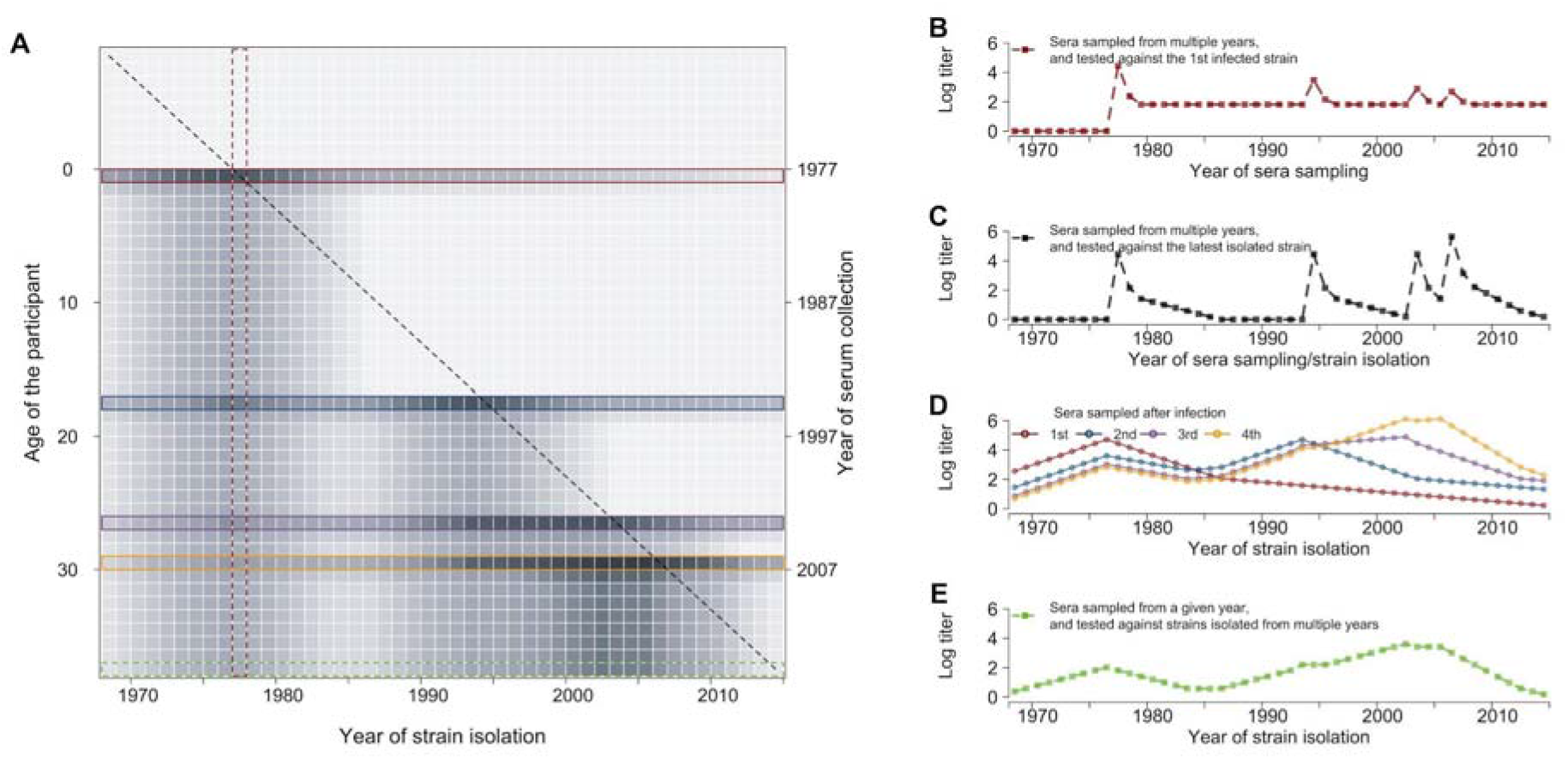
Conceptual plot for individual life-course immune responses to influenza. **(A)** Simulated annual antibody profiles of a representative individual born in 1977 and who had four infections (1977, 1994, 2003 and 2006) (same as in panel D). Each row represents the antibody profile of strains, which were isolated between 1968 and 2014, from serum that was collected on the year listed in the right y-axis and at the participant’s age listed in the left y-axis. Darker grey represents greater HI titer. The diagonal dashed line represents the real-time titer - titer of strain isolated at year t measured at year t -1 that determined the risk of infection in each year. Colored boxes represent antibody profiles that were measured in each year of infection (1st red; 2nd blue; 3rd purple and 4th orange) and sample collection (green, panel E). (B) Antibody responses against the first infected strain that were measured at different years, i.e., the red vertical block in panel A. (C) Antibody responses against the strain that was isolated/circulated in the year of sampling, i.e, the diagonal line in panel A. (D) Antibody profiles of influenza strains at each infection. (E) Antibody profiles of influenza strains at each infection.

**Fig. S4.**
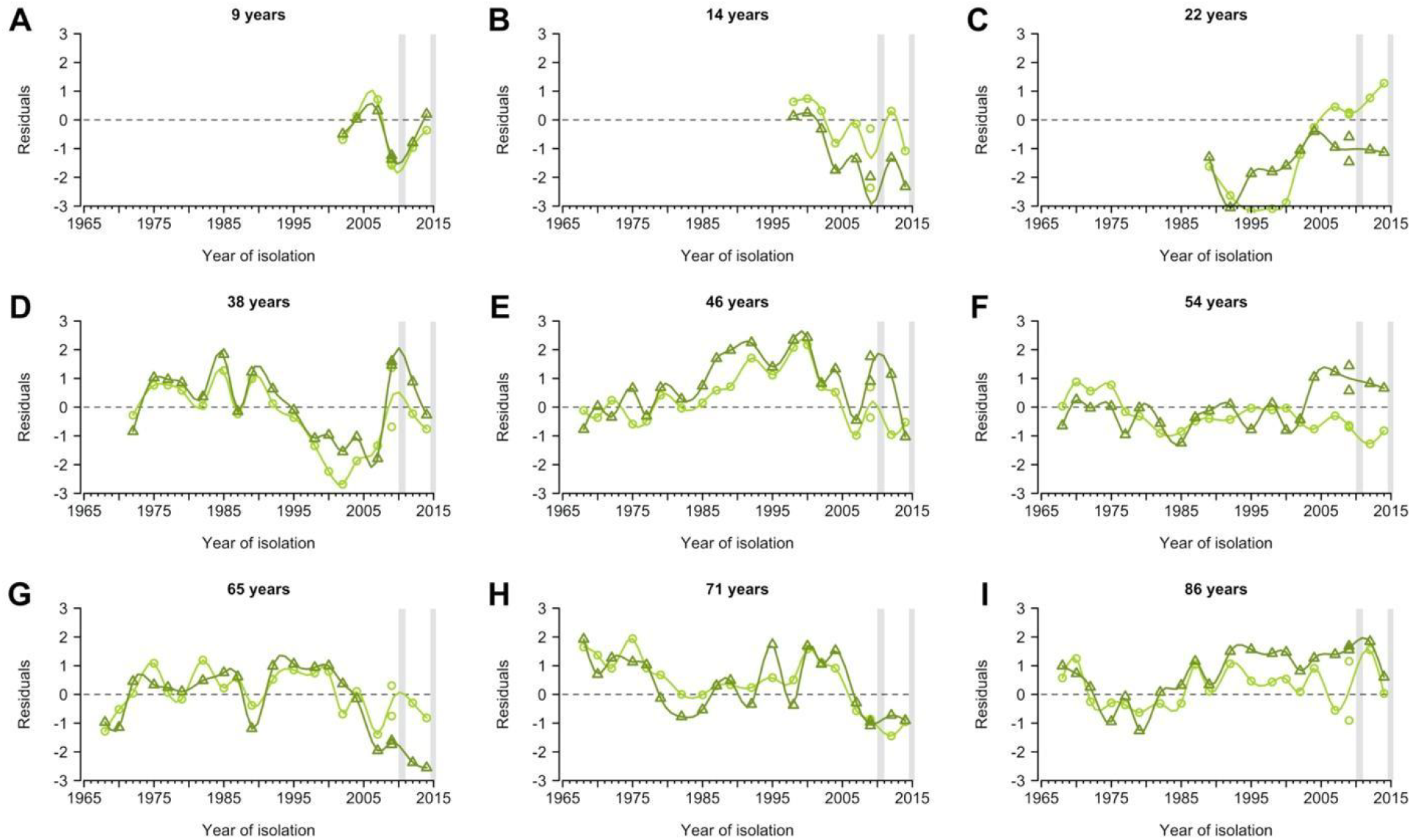
Representative individual profiles of residuals of hemagglutination inhibition (HI) titers. Profiles are from the same individuals shown in Fig. S2. Residuals were derived from fitted generalized additive models of log HI titer on age at sampling, age at circulation and strain (Fig. S3A). Data from the baseline and follow-up visit were fitted separately. Light and dark green represent the baseline and follow-up visit, respectively. Lines were derived from the smooth spline of the observed data points and are shown for illustration purposes only. Grey vertical lines represent the duration of the two visits.

**Fig. S5.**
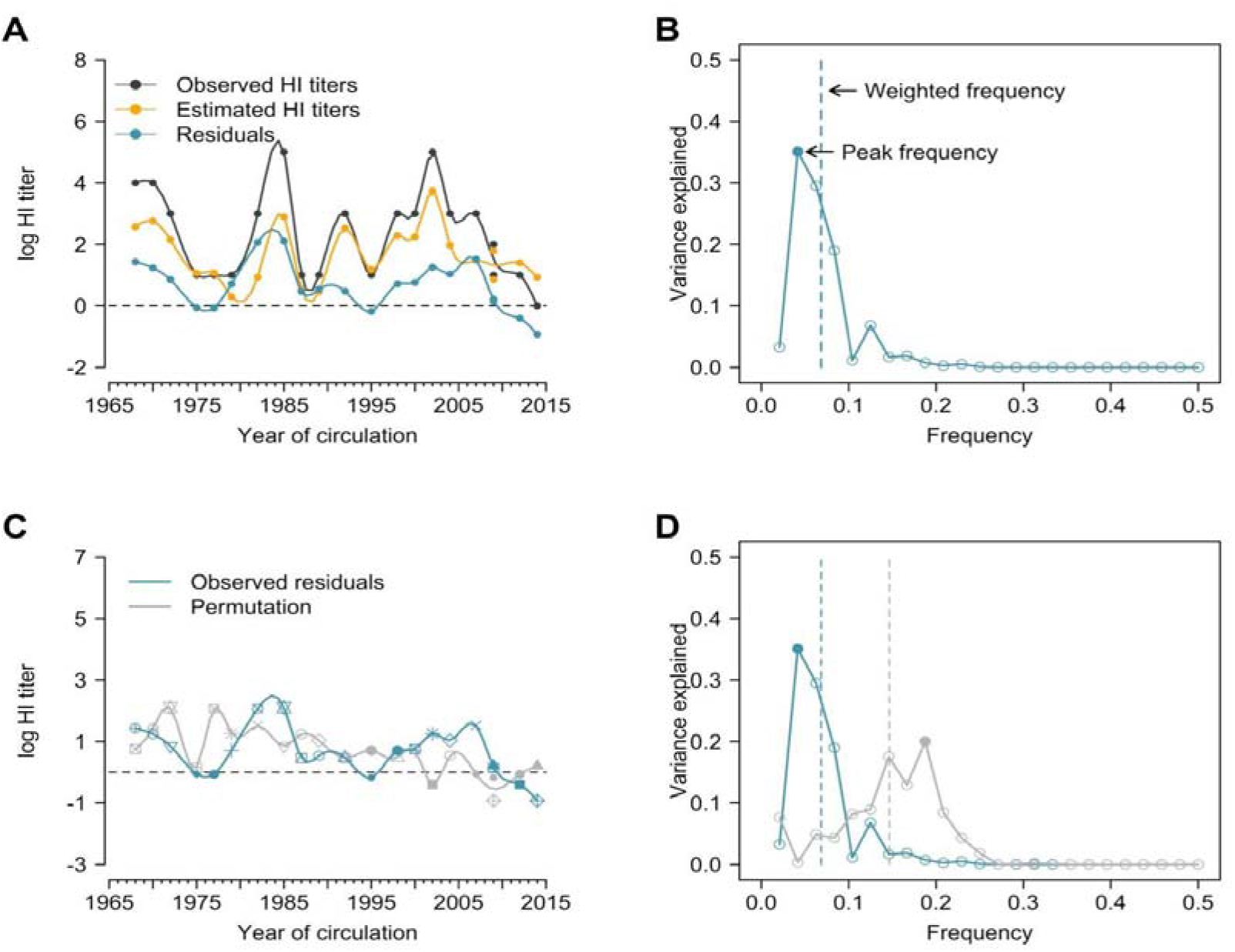
Illustration of estimation of individual time series of residuals and Fourier analysis of observed and permutation of time series. **(A)** Observed HI titers, estimated HI titers and residuals. Estimations were derived from the generalized additive model (GAM) of log HI titers on age at sampling (spline) and age at circulation (spline) with strain-specific intercept. Residuals were calculated as the difference between observed and estimated HI titers (i.e., black minus orange; shown as the blue line). (B) Peak and weighted frequency of a Fourier spectrum of the time series of residuals shown in A. **(C)** Permutation of the time series of residuals. Shapes of points are used to match the observed and permutation of the strains, e.g., the residual value indicated by solid square is for the A/Texas/2012 but was assigned to A/Fujian/2002 for this permutation. (D) Peak and weighted frequencies of the Fourier spectra for the observed and permuted time series of residuals shown in C. Solid points are peak frequencies and dashed lines are weighted frequencies.

**Fig. S6.**
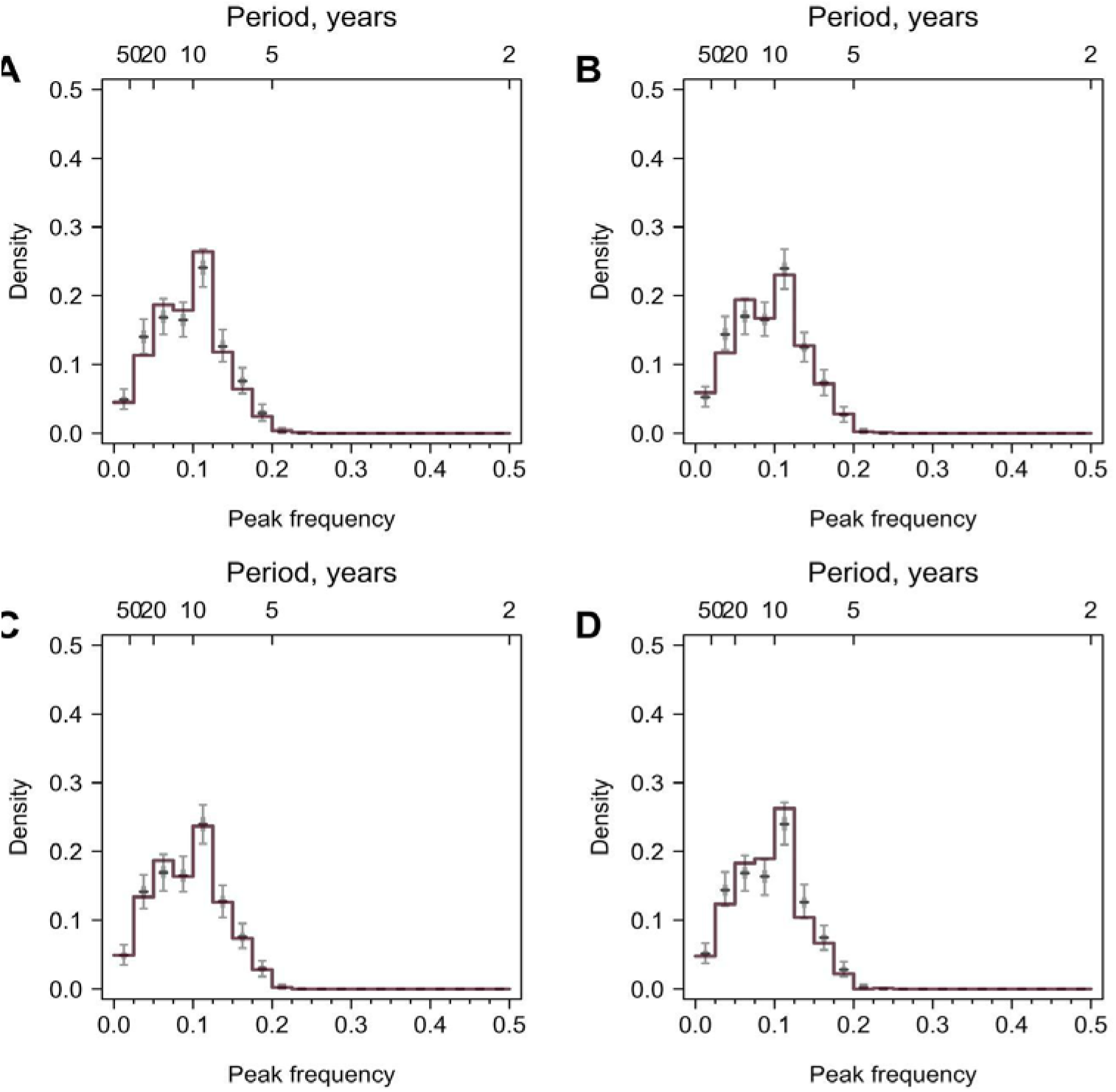
Validation using values generated from random distributions with no periodicity. 777 time series were simulated, and the lengths and resolutions of the time series were the same as the data. Colored lines are distributions of peak frequencies for yearly interpolation of the simulated data, and grey boxplots are distributions for 1,000 permutations of the simulated data. **(A)** Normal distribution. Random values were generated from a normal distribution with mean 1 and sd 2. **(B)** Lognormal distribution. Random values were generated from lognormal distribution with mean 1.5 and sd 0.5. **(C)** Normal distribution with outliers. Random values were generated from the same normal distribution in panel A, in which 2 to 4 time points for each time series had values from a normal distribution with larger mean (mean = 3 and sd =1). **(D)** Lognormal distribution with outliers. Random values were generated from the same lognormal distribution in panel B, in which 2 to 4 time points for each time series had values from a lognormal distribution with larger mean (mean = 2 and sd =0.2).

**Fig. S7.**
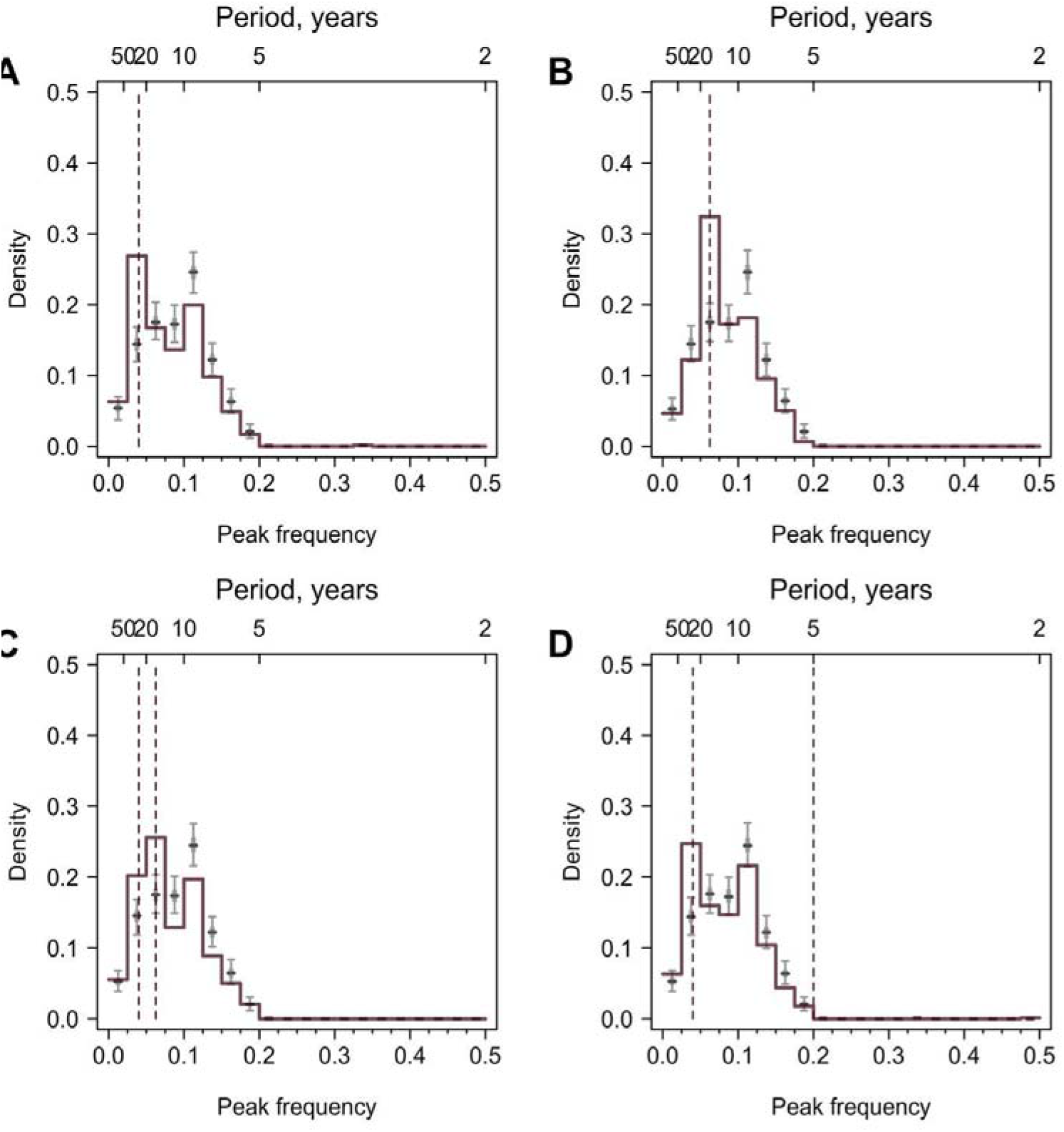
Validation using values generated from periodic curves. A sine curve with predefined periodicity and white noise was simulated on a yearly basis, and values for the time points when tested A(H3N2) strains were isolated were extracted. 777 time series were simulated, and the lengths and resolutions of the time series were the same as the observed data. Colored lines are distributions of peak frequencies for simulated data, and grey lines are distributions for 1,000 permutations of the simulated data. Dashed lines represent the true periodicity. **(A)** All participants have a 25-year periodicity. **(B)** All participants have a 16-year periodicity. **(C)** Half of the participants have a 25-year periodicity and the other half have a 16-year periodicity. **(D)** All participants have periodicities of 25 and 5 years.

**Fig. S8.**
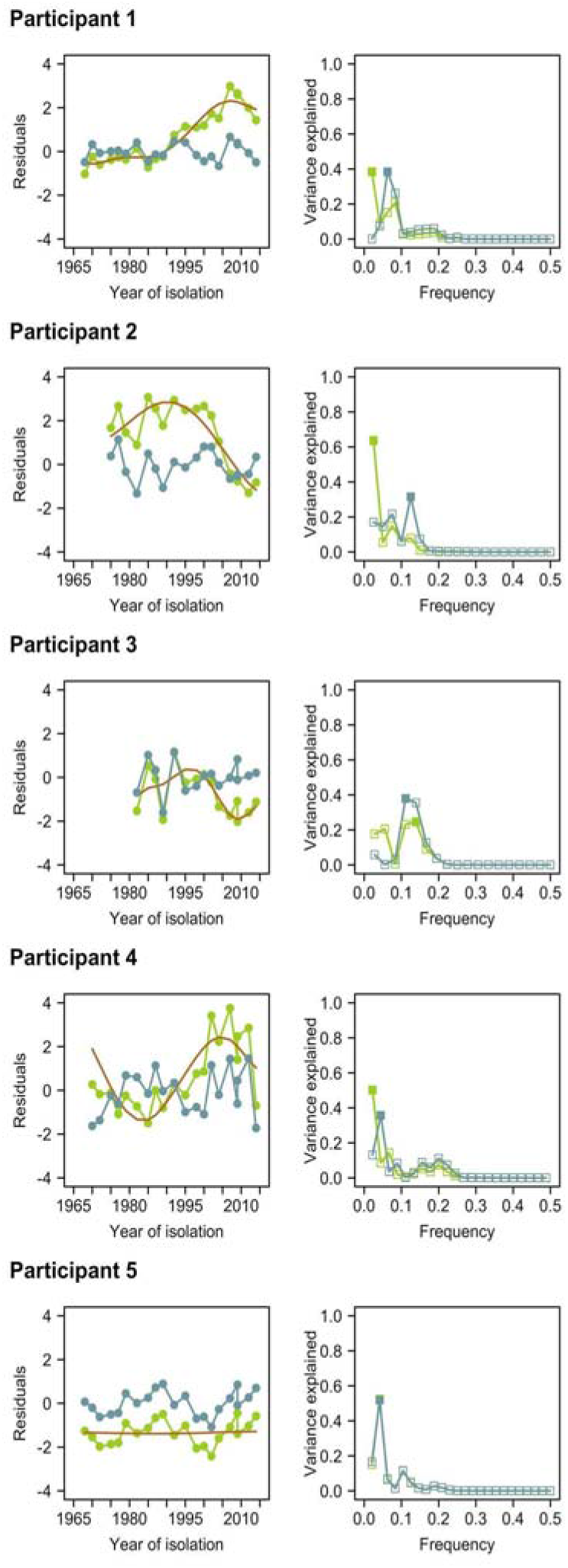
Comparison of time series of residuals and their Fourier spectra before and after empirical mode decomposition (EMD). Left columns represent time series of residuals before (green) and after (blue) detrending using EMD. The orange line indicates the non-linear trend identified using EMD. Right columns represent the Fourier spectra before (green) and after (blue) detrending with EMD. Solid squares indicate the peak frequencies in the Fourier spectra.

**Fig. S9.**
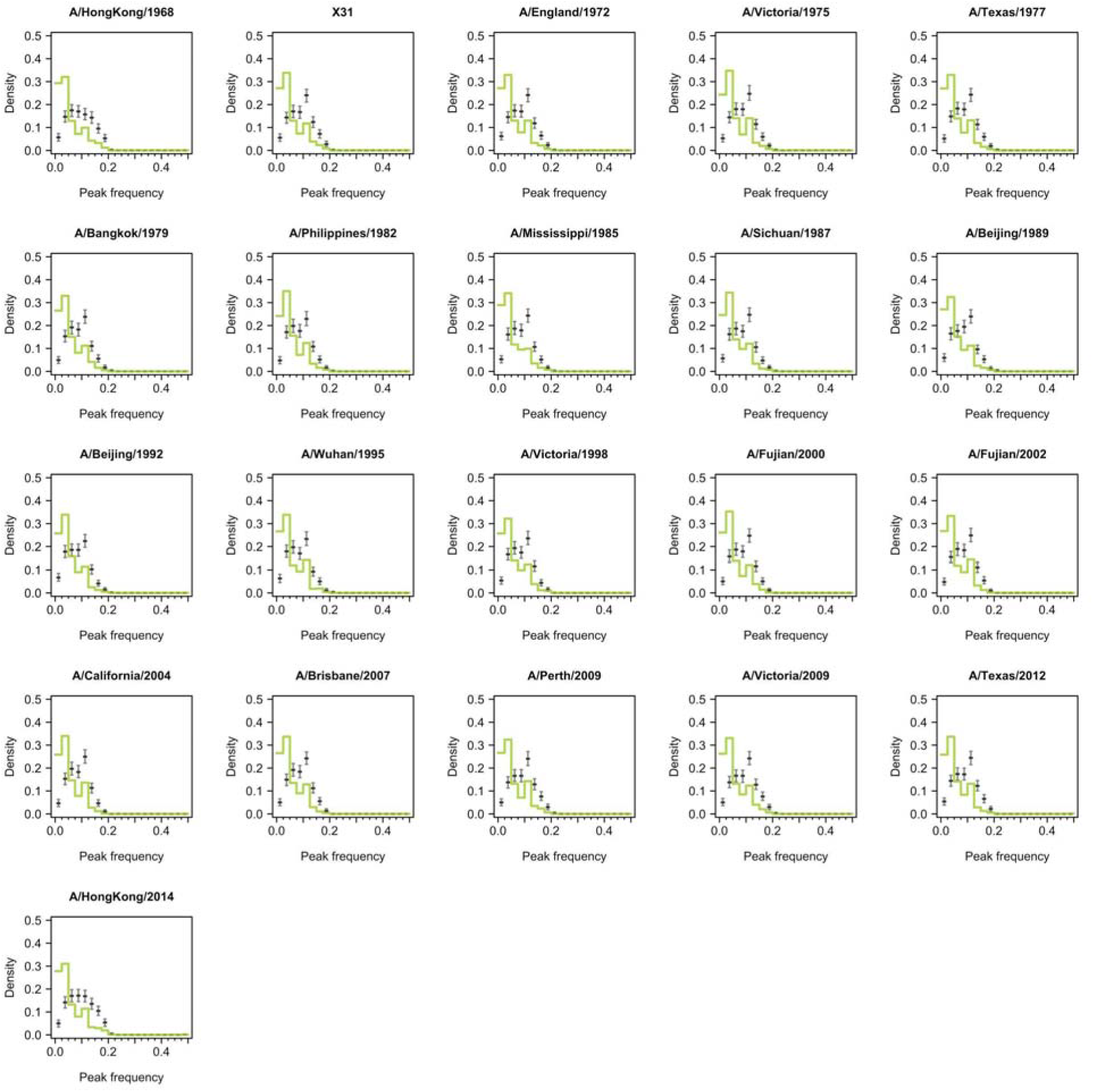
Distribution of peak frequencies across individuals when dropping titers of one tested strain for serums collected at baseline. Light green lines indicate the distribution of peak frequencies in the data. Median (thick grey ticks), interquartile (grey bars) and 95% intervals (think grey ticks) of Fourier spectra from 1,000 permuted time series are indicated.

**Fig. S10.**
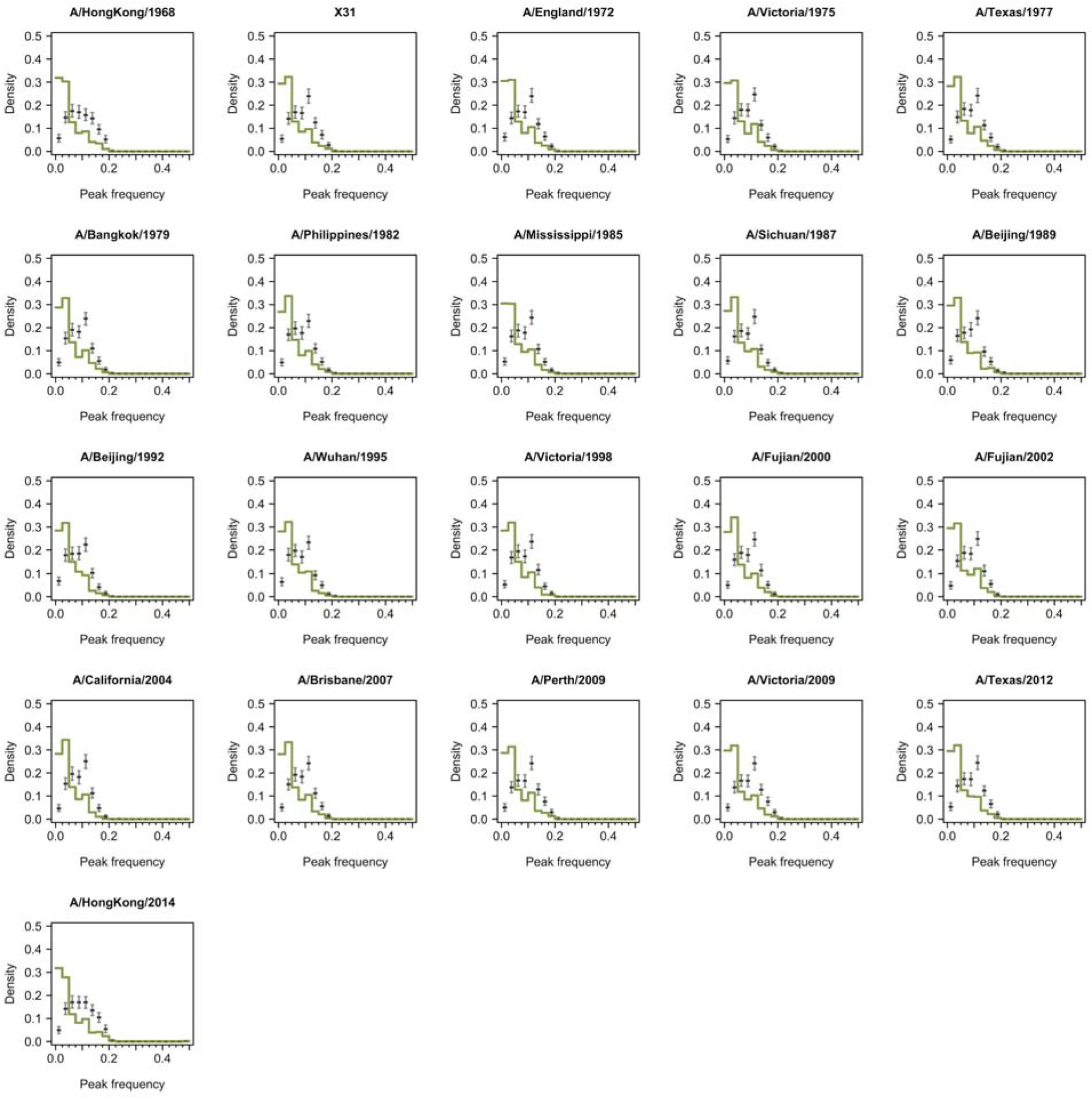
Distribution of peak frequencies across individuals when dropping titers of one tested strain for serums collected at follow-up. Dark green lines indicate the distribution of peak frequencies in the data. Median (thick grey ticks), interquartile (grey bars) and 95% intervals (think grey ticks) of Fourier spectra from 1,000 permuted time series are indicated.

**Fig. S11.**
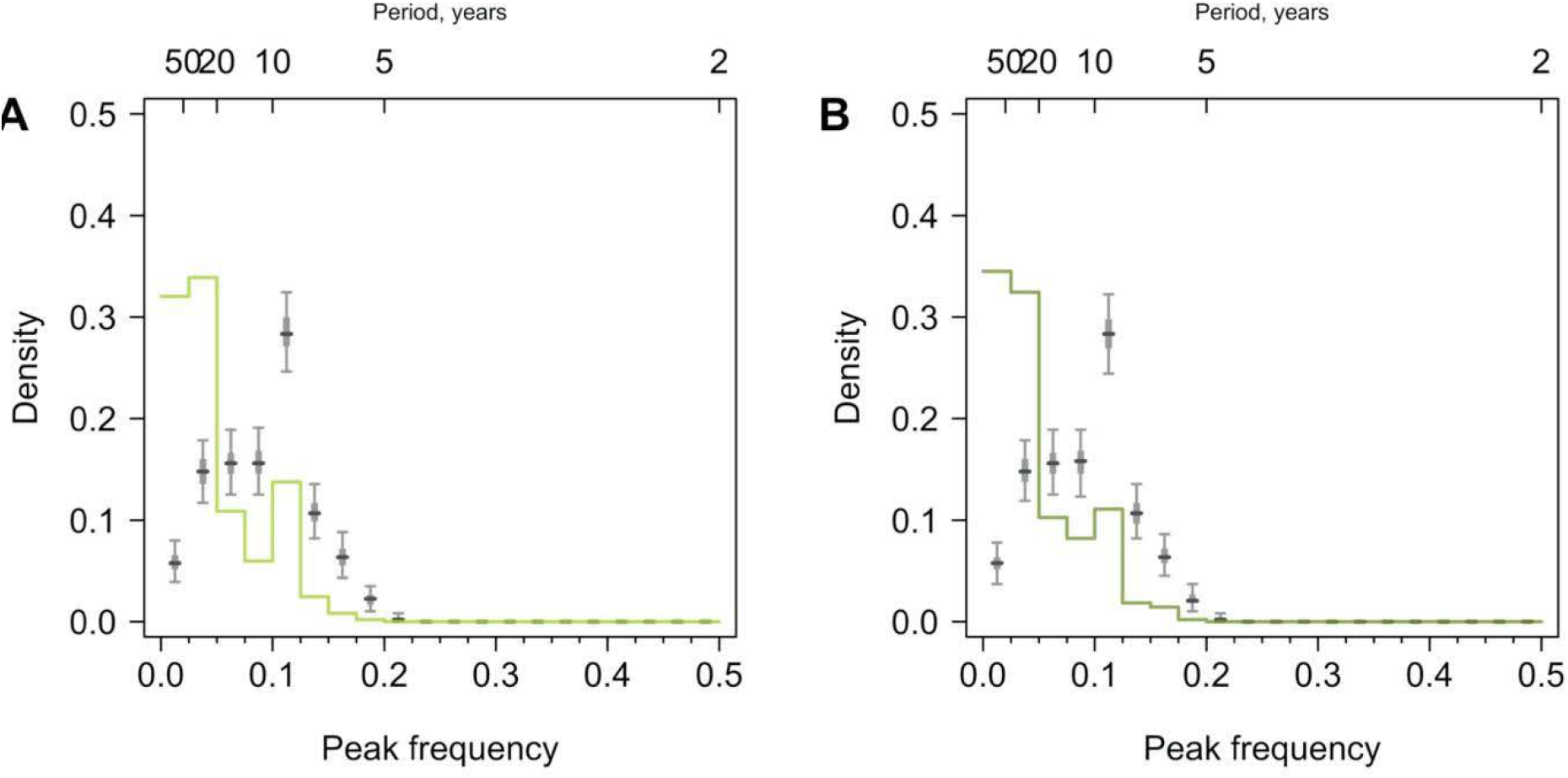
Distribution of peak frequencies across individuals born before 1968. **(A)** Serums collected at baseline. **(B)** Serums collected in the follow-up visit. We performed Fourier spectral analysis on the time series of residuals and extracted peak frequencies for a subset of participants born before 1968 and thus able to experience all tested H3N2 strains. Light and dark green lines represent the distribution of peak frequencies across participants for the baseline and follow-up visits, respectively. Median (thick grey ticks), interquartile (grey bars) and 95% intervals (think grey ticks) of Fourier spectra from 1,000 permuted time series are indicated.

**Fig. S12.**
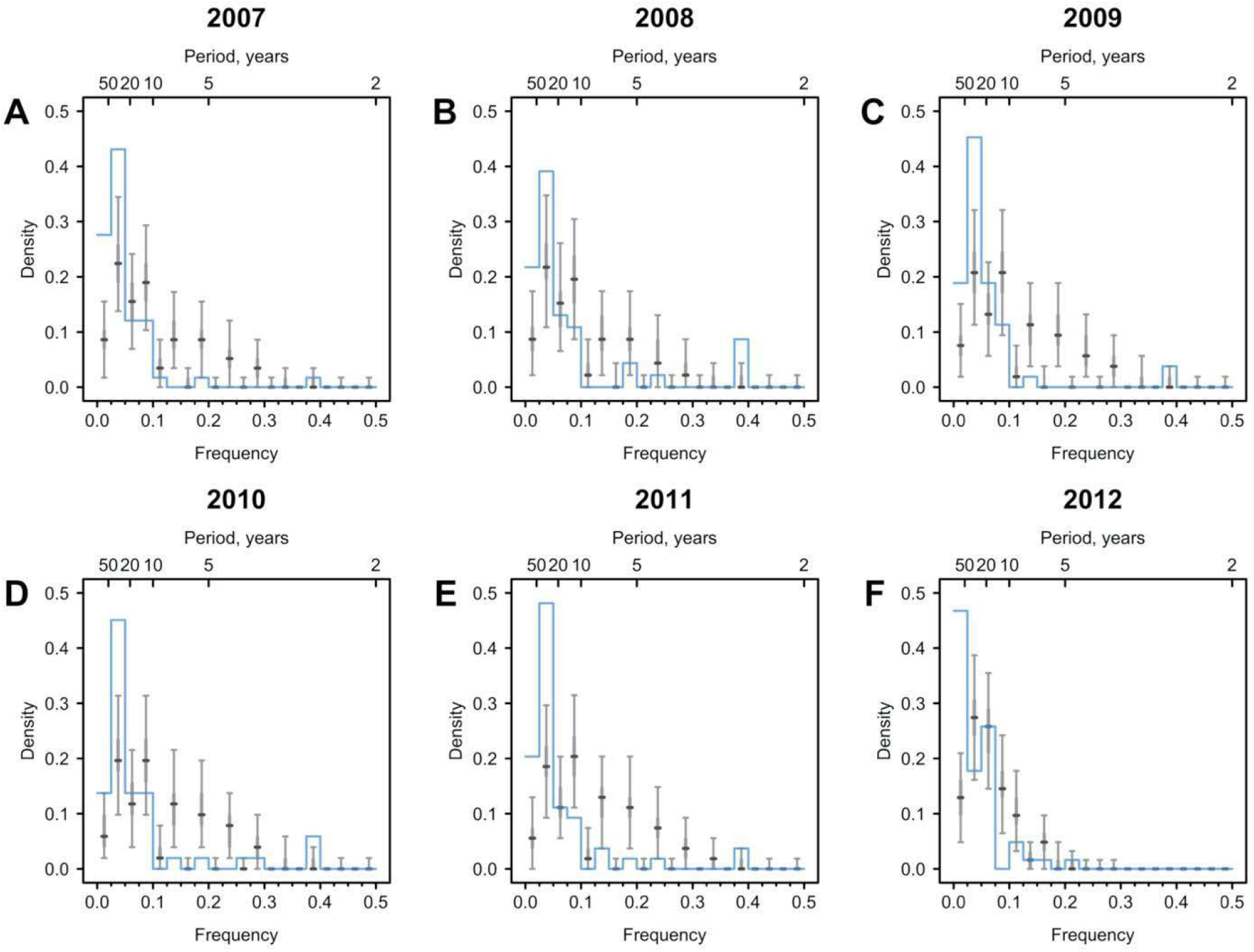
Cycles in immune responses to influenza in the Vietnam data. **a** to **f** show distributions of frequencies that explain the most variance across 69 individuals, whose serums were annually collected from 2007 to 2012. The Vietnam data was taken from (4) as derived from (3, 20). Blue lines are distributions of peak frequencies detected in the observations. Median (thick grey ticks), interquartile (grey bars) and 95% intervals (think grey ticks) of Fourier spectra from 1,000 permuted time series are indicated.

**Fig. S13.**
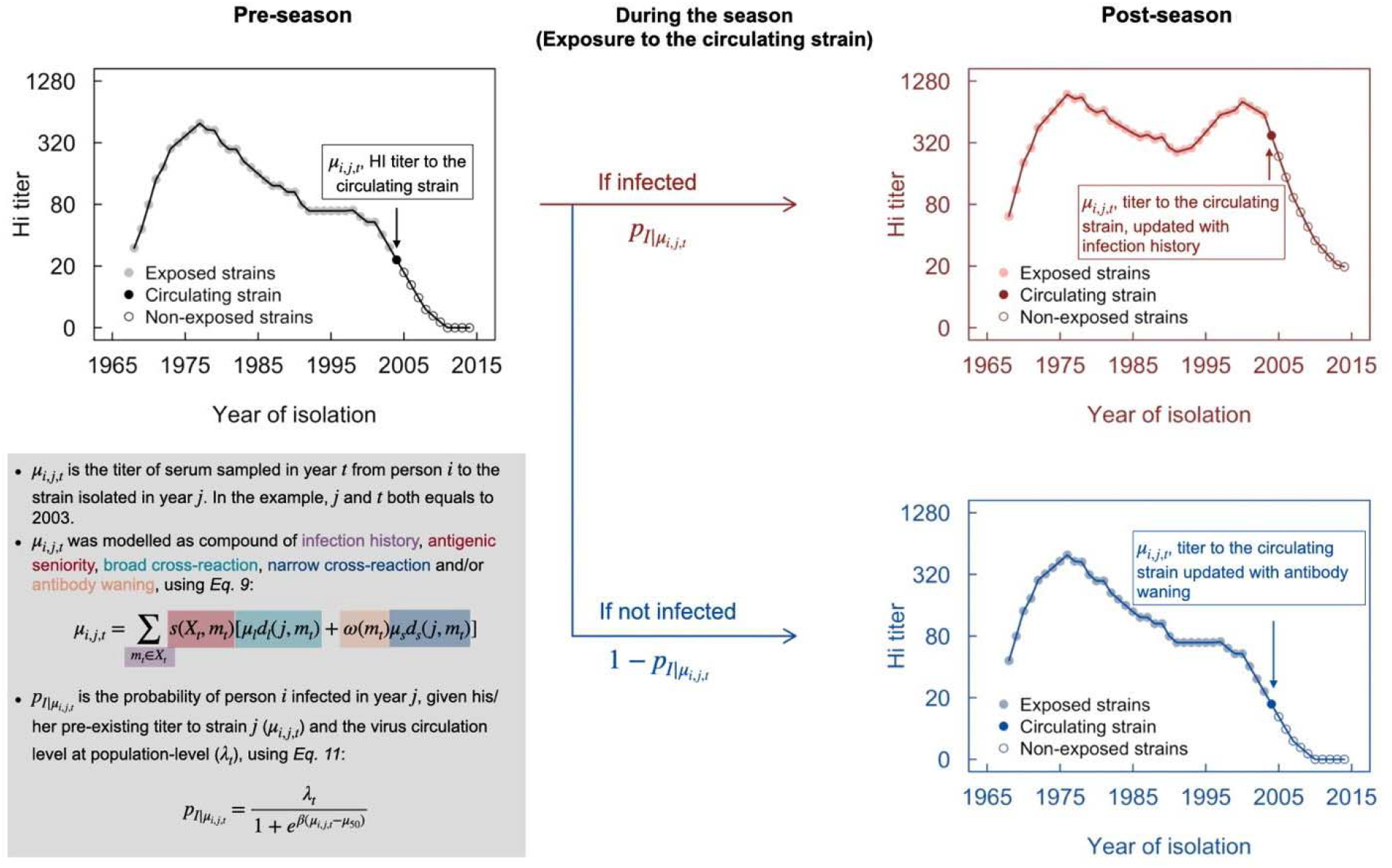
Conceptual plot of modeling immune responses. The black line in the top-left panel denotes pre-season (e.g., 2003) antibody profile, from which the titer of the circulating strain (filled point) was used to determine the probability of infection, using *Equation 11*. Infection was randomly generated following a binomial distribution with probability p. Red and blue lines in the right panels indicate the post-season antibody profile for infection and non-infection, respectively.

**Fig. S14.**
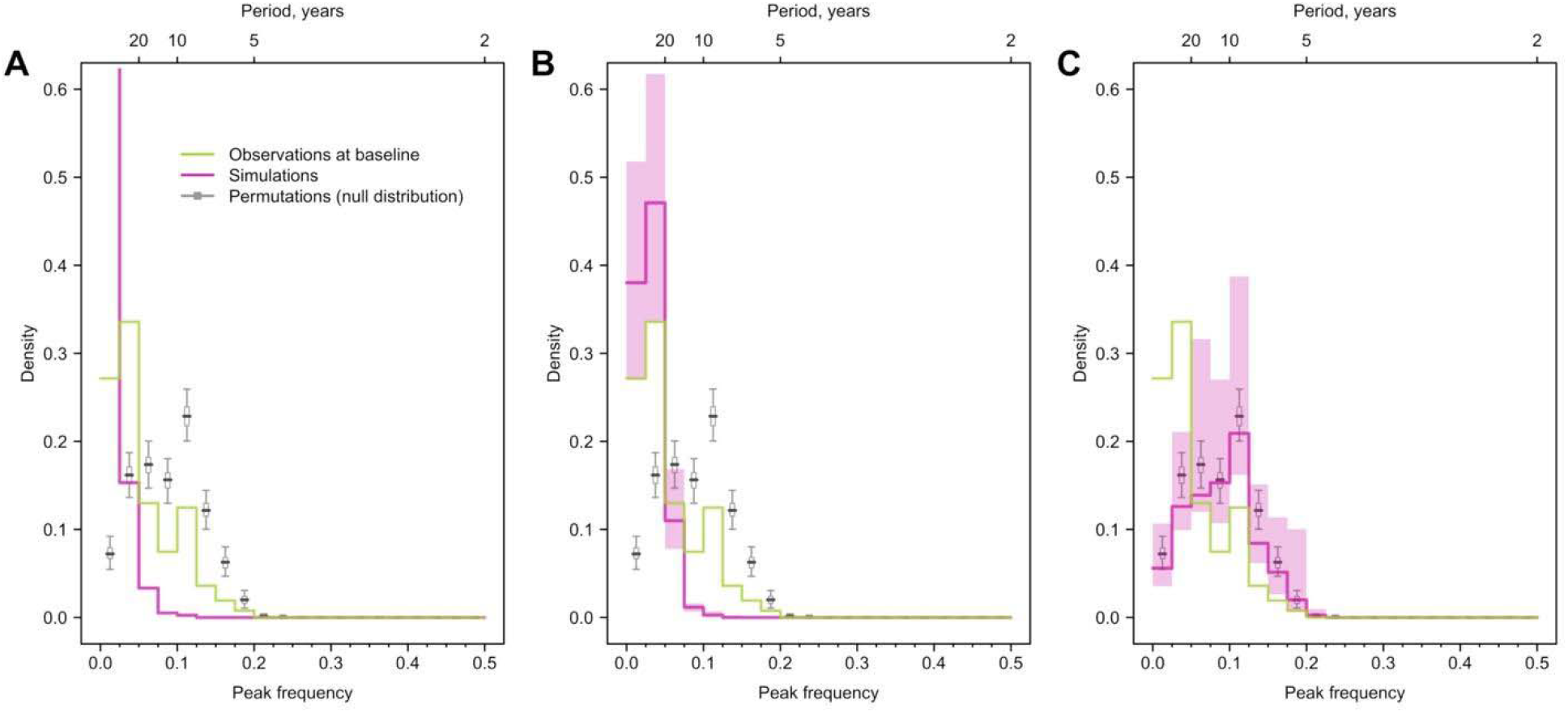
Impact of antigenic evolution speed on the reported cycles in individual antibody responses. Green and purple lines indicate the distribution of peak frequencies detected in the observed simulated antibody profiles across individuals. Grey lines are the distributions of peak frequencies of the 1,000 permutations of the simulated antibody profiles (i.e., null distribution). We assumed the antigenic evolution speed was 5 times slower **(A)**, the same **(B)** and 5 times faster **(C)** than that was used in the main analysis.

